# Human cell-camouflaged nanomagnetic scavengers restore immune homeostasis in a rodent model with bacteremia

**DOI:** 10.1101/2022.03.16.22272481

**Authors:** Sung Jin Park, Seyong Kwon, Min Seok Lee, Bong Hwan Jang, Axel E. Guzmán-Cedillo, Joo H. Kang

## Abstract

Bloodstream infection caused by antimicrobial resistance pathogens is a global concern because it is difficult to treat with conventional therapy. Here we report scavenger magnetic nanoparticles enveloped by nanovesicles derived from blood cells (MNVs), which magnetically eradicate an extreme range of pathogens in an extracorporeal circuit. We quantitatively reveal that glycophorin A and complement receptor (CR) 1 on red blood cell (RBC)-MNVs predominantly capture human fecal bacteria, carbapenem-resistant (CR) *E. coli*, and extended-spectrum beta-lactamases-positive (ESBL-positive) *E. coli*, vancomycin-intermediate *S. aureus* (VISA), endotoxins, and proinflammatory cytokines in human blood. Additionally, CR3 and CR1 on white blood cell-MNVs mainly contribute to depleting the virus envelope proteins of Zika, SARS-CoV-2, and their variants in human blood. Supplementing opsonins into the blood significantly augments the pathogen removal efficiency due to its combinatorial interactions between pathogens and CR1 and CR3 on MNVs. The extracorporeal blood cleansing enables full recovery of lethally infected rodent animals within seven days by treating them twice in series. We also validate that parameters reflecting immune homeostasis, such as blood cell counts, cytokine levels, and transcriptomics changes, are restored in blood of the fatally infected rats after treatment.

## 1. Introduction

Infectious diseases persistently threaten a large population worldwide, resulting in 11 million deaths and 49 million cases annually.^[1]^ The introduction of antibiotics^[2]^ and antiviral drugs^[3]^ has greatly mitigated the social and economic burdens of infectious diseases; however, the unpredictable appearance of a new strain of pathogens recalcitrant to existing drugs catastrophically impacts the public. Because a new strain of bacteria has emerged that is resistant to existing antibiotics^[4]^, an improved therapeutic strategy is needed to treat patients infected with pandrug-resistant bacteria (PDR). The incidence of infection caused by antibiotic-resistant bacteria became one of the greatest concerns for forthcoming generations because antimicrobial resistance is predicted to be the leading cause of death in 2050.^[5]^ However, the therapeutic options for the recent increases in the prevalence of carbapenem-resistant *Enterobacteriaceae* (CRE), carbapenem-resistant *Acinetobacter baumannii* (CRAB), and carbapenem-resistant *Pseudomonas aeruginosa* (CRPA) are currently limited.^[6-8]^ Moreover, for neonatal blood infections, antibiotic use is known to result in adverse life-long effects on the microbiome,^[9,10]^ potentially inducing disease-promoting gut microbiota alteration.^[11]^ More importantly, because of the unculturable bacteria in bloodstream infection (BSI),^[12]^ the frequent false negative blood culture outcomes (∼70%),^[13]^ and the unavailable diagnostic tools for detecting endotoxin levels in whole blood,^[14]^ removing both detectable and undetectable pathogenic materials in the bloodstream is especially critical.

In the pandemic era, despite the extensive efforts that have been made to develop a new antibiotic drug or antiviral vaccine throughout the world, drugs and vaccines are often elusive due to the long process necessary (∼10 years) for developing a new drug candidate^[15,16]^; throughout the process, the public must withstand the pressure caused by the outbreak as there is no proper therapeutic strategy. Because the viral loads in blood reflect the mortality of patients infected with SARS-CoV-2,^[17]^ a new therapeutic strategy for simultaneously depleting both viruses and proinflammatory cytokines in the patient’s blood should also be considered.

As a resource interacting with all organs throughout the body, blood is responsible for spreading inflammatory cascades via the immune and cardiovascular systems,^[18]^ which could lead to multiple organ dysfunction and deaths regardless of the infection site.^[19]^ Many previous efforts validated that reducing pathogenic loads in blood using extracorporeal devices could alleviate the clinical outcomes of patients with bacteremia or endotoxemia.^[20,21]^ However, there are considerable concerns that the therapeutic efficacy of extracorporeal blood treatment has not been completely validated. One factor that contributes the inconclusive nature of these results may be the lack of tools that can effectively eradicate a wide range of pathogens and cytokines while simultaneously contributing to systemic inflammatory responses; this lack of tools may be because capturing reagents require a high mobility for the sequestration of pathogenic materials from blood^[22,23]^ or due to the inability of binding the intact clinical pathogens found in patients.^[24]^

To address the unmet challenges in conventional extracorporeal blood treatment, we developed magnetic nanovesicles (MNVs) covered with blood cell membranes, such as human red blood cells (hRBCs) and human white blood cells (hWBCs), which are known to inherently bind a broad range of pathogens, including bacteria, viruses, and endotoxins. Cell membrane-derived nanoparticles have been used for targeted drug delivery,^[25]^ cancer therapy, ^[26]^ and even for treating neutralizing infectious reagents^[27,28]^ and capturing clinically important bacterial species.^[29]^ However, no efforts have been made to explore the daunting task of simultaneously removing various pathogenic materials, which would result in immense therapeutic efficacy. Our approach is unique because it allows us to physically remove a broad range of both pathogenic materials and proinflammatory cytokines from whole blood without triggering the undesirable immune reactions, which are potentially caused by magnetic nanoparticles during incubation with blood, because their surfaces are camouflaged with human cell membranes. Our approach offers a new therapeutic strategy that simultaneously targets a broad range of clinical isolates of various bacteria, viruses, and proinflammatory cytokines for the first time.

## 2. Result and discussion

### 2.1 Cell membrane-derived magnetic nanovesicles and blood-cleansing system

MNVs were designed to be capable of simultaneously binding an extensive range of pathogens, endotoxins, and proinflammatory cytokines, all of which consistently trigger sepsis cascades. We aimed to develop MNVs that indirectly or directly capture a range of pathogens with or without various plasma opsonin molecules to maximize the binding diversity of MNVs. Among the plasma membrane proteins involved in capturing pathogens in human blood, glycophorin A (GYPA), complement receptors 1 (CR1), and CR3 were considered the most promising candidates because GYPA, the most abundant sialoglycoprotein on RBCs, directly binds to diverse pathogens and their toxins^[30]^; CR1 interacts with multiple opsonins, resulting in diverse pathogen-binding mechanisms, such as C1q, C3b, iC3b, collectins, and ficolins^[31]^; and CR3 has been shown to bind pathogens directly or indirectly using its lectin-like site or via iC3b^[32]^. Moreover, to capture proinflammatory cytokines simultaneously along with pathogens using MNVs, the Duffy antigen receptor for chemokines (DARC) present on hRBCs^[33]^ and diverse cytokine binding receptors on hWBCs^[34]^ were also explored for the treatment of the patients with infectious diseases by removing those pathogenic materials from the circulating blood (**Figure 1**a,b). We then discovered that hRBCs and hWBCs are human cells that simultaneously possess all the plasma membrane proteins we intended to incorporate into the MNV surfaces. To validate the superiority of the pathogen-binding efficiency of hRBC-MNVs and hWBC-MNVs, we also generated MNVs with other cells or blood components, including human hepatic sinusoidal cells (hHSECs), U937 cells, HL60 cells, and platelets, that are known to be involved in immunological activity.

**Figure 1.**
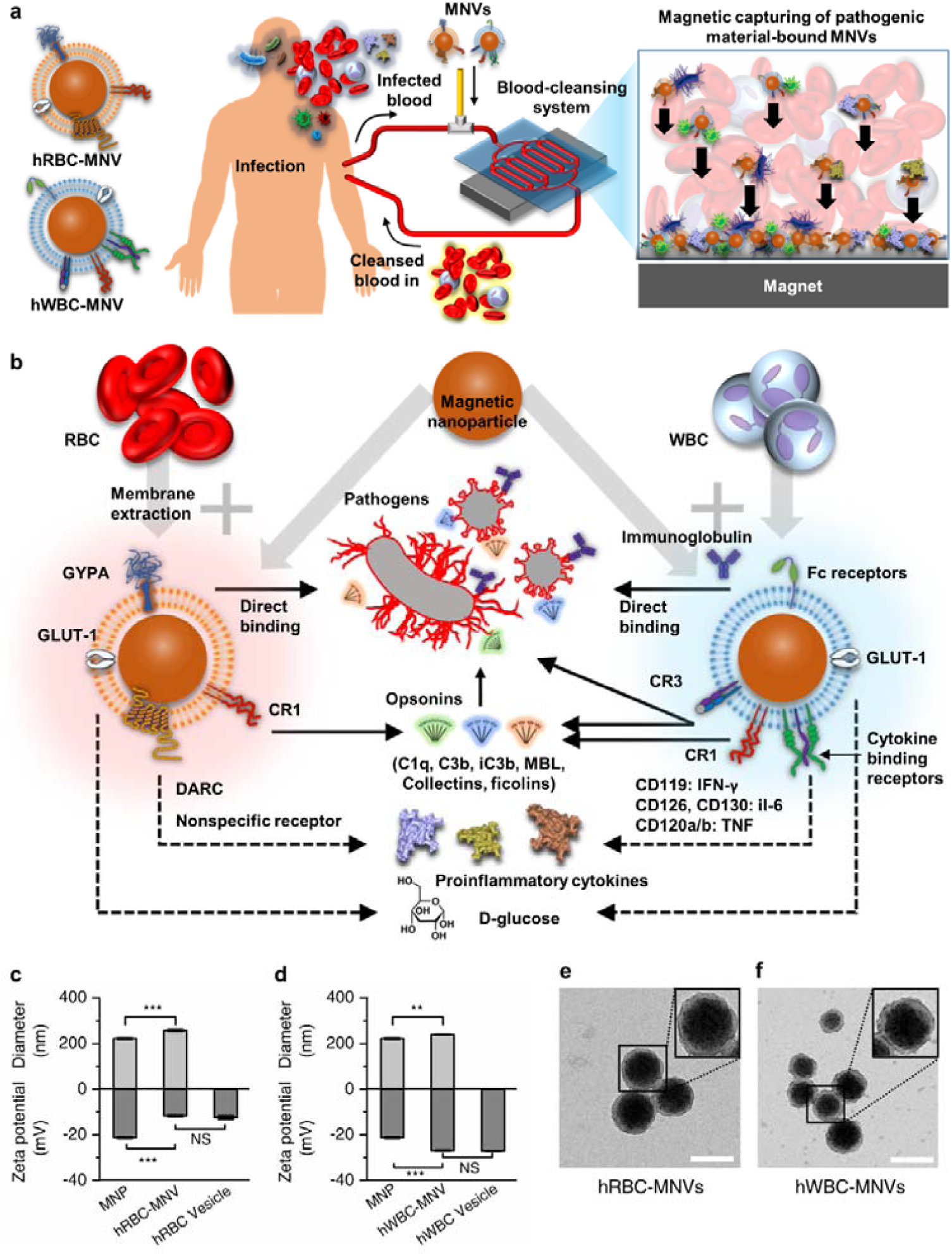
Magnetic nanovesicles (MNVs) and magnetic blood-cleansing system. a) Magnetic nanoparticles (MNPs) covered with blood cell membranes (RBCs and WBCs) represent the intrinsic surface receptors responsible for capturing a range of pathogens and proinflammatory cytokines in the blood, including glycophorin A (GYPA), Duffy antigen receptor for cytokines (DARC), CR1, CR3, Fc receptors, and various cytokine-binding receptors. GYPA on RBC-MNVs directly binds to various bacteria and viruses by functioning as a decoy to pathogens. CR1 and CR3 bind to a broad range of pathogens via opsonin molecules such as C3b, iC3b, C1q, collectins, and ficolins, whereas Fc receptors on WBC-MNVs engage a specific pathogen recognized by pathogen-specific immunoglobulins with a high affinity. DARC on RBC-MNVs and cytokine-binding receptors on WBC-MNVs, including CD119, CD126, CD130, and CD120a/b, capture proinflammatory cytokines in the blood. b) A schematic of the blood cleansing treatment for the patient with infectious diseases using MNVs. c,d) The hydrodynamic diameter and zeta potential of the fabricated MNPs, membrane vesicles without MNP cores, and MNVs made of c) hRBCs and d) hWBCs (*n* = 3). e,f) Transmission electron microscopy images of e) hRBC-MNVs and f) hWBC-MNVs. Scale bar, 200 nm. Data were expressed as means ± SEM. Statistical significance was calculated by a two-tailed Student’s t test. \*\**P* < 0.005; \*\*\**P* < 0.001; NS, not significant.

To develop MNVs, we encapsulated magnetic nanoparticles (MNPs) in nanovesicles derived from hRBCs and hWBCs by serial extrusions of ultrasonicated cell membranes and MNPs sequentially through 1 µm, 0.4 µm, and 0.2 µm pore membranes^[35]^. The hydrodynamic diameter of MNPs increased from 220 ± 3.96 nm to 258 ± 4.69 nm after the MNVs were formed, and their surface zeta potential became comparable to those of the cell membrane nanovesicles without MNP cores, supporting that the MNPs were completely encapsulated with the cell membranes (Figure 1c,d and Figure S1). Transmission electron microscopy (TEM) images of hRBC-MNVs and hWBC-MNVs show core-shell structures, verifying that the cell membrane was successfully translocated onto the MNP surface (Figure 1e,f).

The expanded binding capability of the MNVs encapsulated with various human cells was validated using methicillin-resistant *Staphylococcus aureus* (MRSA), extended-spectrum β-lactamase (ESBL)-positive *Escherichia coli*, respiratory syncytial virus (RSV), cytomegalovirus (CMV), Zika virus E protein, and SARS-CoV-2 S protein, which present the most daunting challenges among global infectious diseases. Among those MNVs, the hRBC-MNVs yielded the best affinity to bacteria in human whole blood (Figure S2a,b), and the hWBCs-MNVs exhibited the most proficient binding efficiency to those viruses or virus envelope proteins, despite the presence of CR1 and CR3 on U937 and HL60, respectively; scavenger receptors and mannose receptors in hHSEC; and GPIIb–IIIa, GPIbα, gC1q-R, and toll-like receptors in platelet (Figure S2c-f). This is likely due to the fewer molecules of CR1 and CR3 in HL60 and U937 than in hWBCs; the pathogen-binding capacity of scavenger receptors and mannose receptors in hHSECs is competitively inhibited for a wide range of waste materials, such as lipoproteins,^[36]^ lysosomal enzymes,^[37]^ and collagen molecules^[38]^; and without activation, pathogen-binding receptors in platelets have a low binding affinity to bacteria.^[39]^

These results correspond to previous studies reporting that RBCs bind numerous pathogens^[40]^ and the subpopulation of WBCs capture a range of viruses.^[41-46]^ Thus, hRBC-MNVs and hWBC-MNVs were exploited in our extracorporeal blood-cleansing device to achieve the depletion of a broad spectrum of bacteria and viruses, respectively.

We performed quantitative analysis of the hRBC- and hWBC-MNV surface molecules responsible for capturing a range of pathogens and proinflammatory cytokines. hRBC-MNVs retained GYPA, CR1, and DARC, while hWBC-MNVs contained CR1 and CR3 (**Figure 2**a), as we intended.^[47,48]^ We then quantitatively discriminated the contributory surface receptor to capture a range of pathogens, including MRSA, ESBL-positive *E. coli*, RSV, CMV, ZIKV E protein, HCoV-OC43, and SARS-CoV-2, by inactivating each receptor with their corresponding antibody. Both GYPA and CR1 on hRBC-MNVs were determined to play a critical role in capturing bacteria, while direct binding via GYPA did not significantly deplete viruses except for CMV (Figure 2b). The viruses were effectively captured by opsonins in blood, which were then bound to CR1 on hRBC-MNVs (Figure 2b). It was determined that, compared to CR1, CR3 had a major role in the depletion of both bacteria and viruses on hWBC-MNVs, which was most likely due to the higher concentration of CR3 on hWBC-MNVs and their dual pathways that target pathogens either directly or via opsonin molecules (Figure 2c). Additional surface receptors, such as the Fc receptor, Toll-like receptors,^[49]^ and natural cytotoxicity receptors,^[50]^ contributed to capturing pathogens because blocking both CR1 and CR3 did not completely nullify the binding capability of hWBC-MNVs to the pathogens.

**Figure 2.**
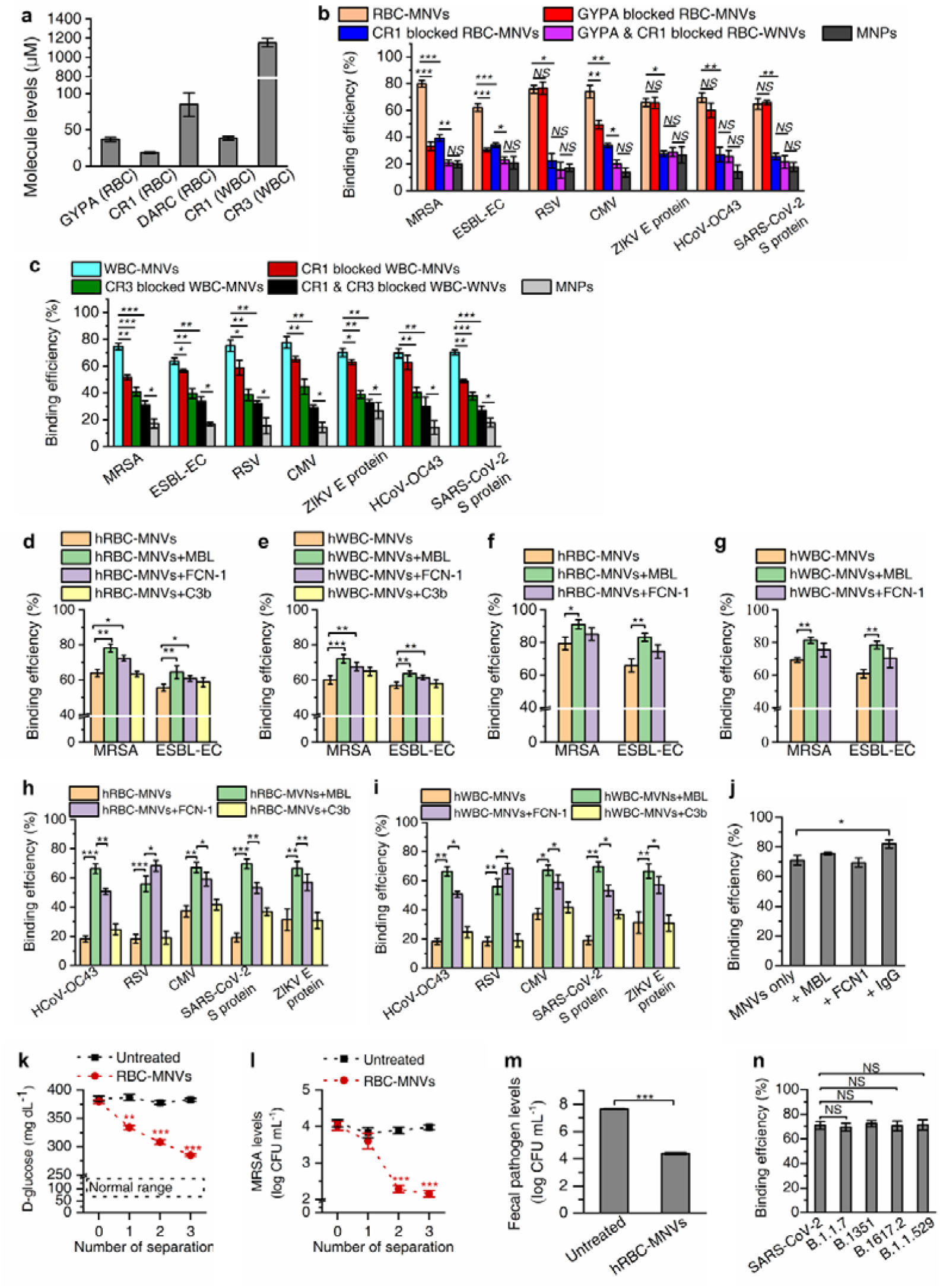
Characterization and pathogen-binding efficiency of MNVs. a) Quantitative measurement of the pathogen-binding-related surface receptors on hRBC-MNVs and hWBC-MNVs. b) Binding efficiencies of hRBC-MNVs, GYPA-blocked hRBC-MNVs, CR1-blocked hRBC-MNVs, GYPA- and CR1-blocked hRBC-MNVs to methicillin-resistant *S. aureus* (MRSA), extended-spectrum β-lactamase-positive *E. coli* (ESBL-EC), RSV, CMV, Zika virus (ZIKV) E protein, HCoV-OC43, and SARS-CoV-2 S protein in human plasma. c) Binding efficiencies of hWBC-MNVs, CR1-blocked hWBC-MNVs, CR3-blocked hWBC-MNVs, and CR1- and CR3-blocked hWBC-MNVs to MRSA, ESBL-EC, RSV, CMV, ZIKV E protein, HCoV-OC43, and SARS-CoV-2 S protein in human plasma. d,e) Binding efficiencies of d) hRBC-MNVs and e) hWBC-MNVs to MRSA and ESBL-EC when supplemented with MBL, ficolin (FCN)-1, or C3b in TBS buffer. f,g) Binding efficiencies of f) hRBC-MNVs and g) hWBC-MNVs to MRSA and ESBL-EC when supplemented with MBL, FCN-1, or C3b in human blood. h,i) Binding efficiency of h) hRBC-MNVs and i) hWBC-MNVs to HCoV-OC43, RSV, CMV, SARS-CoV-2 S protein, and Zika virus (ZIKV) E protein in TBS buffer supplemented with MBL, FCN-1, or C3b. j) Binding efficiency of hWBC-MNVs to SARS-CoV-2 S protein when supplemented with MBL, FCN-1, or anti-SARS-CoV-2 S protein immunoglobulin G (IgG). k) D-glucose depletion in diabetic rat blood by repetitive incubation and magnetic depletion using hRBC-MNVs. l) MRSA spiked in diabetic rat blood was magnetically depleted using hRBC-MNVs. m) Fecal bacterial concentrations in human whole blood were significantly (99.97%) reduced after a single round of magnetic depletion using hRBC-MNVs. n) The removal efficiency of SARS-CoV-2 spike protein and their variants (B.1.1.7, B.1351, B.1.617.2 and B.1.1.529) using hWBC-MNVs. Data were expressed as means ± SEM. Statistical significance was calculated by a two-tailed Student’s t test. \**P* < 0.05; \*\**P* < 0.005; \*\*\**P* < 0.001; NS, not significant.

As we validated the quantitative contributions of CRs for capturing a range of pathogens, we then set out to elucidate which opsonin molecules play a major role in amplifying the pathogen-capturing efficiency of MNVs. We supplemented TBS buffer solutions containing each type of pathogen with the individual opsonin molecules known to interact with CRs, including MBL, ficolin-1, -2, -3, collectin-10, -11, C3b, and C1q. Among these, MBL and ficolin-1 significantly enhanced the binding efficiency of hRBC-MNVs (Figure 2d) and hWBC-MNVs (Figure 2e) to MRSA- and ESBL-positive *E. coli*, supporting the idea that CR1 on hRBC-MNVs and CR1 and -3 on hWBC-MNVs leveraged both MBL and ficolin-1 to capture opsonized bacterial cells. However, in human whole blood, supplementation with MBL only increased the binding efficiency (Figure 2f,g) because ficolin-1 is inherently present in the blood at a low level (∼0.3 µg mL^-1^),^[51-53]^ and thus, replenishing ficolin-1 into the blood does not considerably induce additional bacteria opsonization.

In the context of capturing a wide range of pathogenic viruses using MNVs, MBL and ficolin-1 also played a major role in opsonizing the various viruses, where MBL was more critical in opsonizing the viruses to be captured by CR1 on hRBC-MNVs except for RSV (Figure 2h). Compared to MBL, RSV was more synergized with ficolin-1 because the fibrinogen-like recognition domain of ficolin-1 binds to the mucin-like domain of glycoproteins in RSV with a high affinity.^[54, 55]^ The virus capturing efficiency of the hWBC-MNVs without the added opsonin was superior to that of hRBC-MNVs (Figure 2i) because the CR3 present on the hWBC-MNVs could directly bind to gp41, the viral transmembrane protein.^[56]^ Most importantly, we validated that supplementing virus-targeting IgG in the blood further augmented the capturing efficiency of SARS-CoV-2 spike proteins due to the Fc receptors on hWBC-MNVs (Figure 2j), which reveals that adaptive immunity acquired either by vaccination or prior exposure to the virus could improve the removal efficiency when hWBC-MNVs deplete the viruses within blood.

In addition to the pathogen-binding proteins, hRBC-MNVs and hWBC-MNVs have glucose transporter 1 (GLUT-1), which captures glucose in blood. Patients with diabetes are more vulnerable to infections due to the compromised antibacterial activity of immune cells, such as impaired phagocytosis.^[57,58]^ High glucose levels in the blood are predicted to interfere with the binding of MNVs to pathogens because glucose competes with MBL for binding the carbohydrate domains on pathogens,^[59]^ which then decreases the capturing efficiency of CR1 on MNVs to pathogens via MBL. To resolve this, we leveraged GLUT-1 on hRBC-MNVs, which could lower blood glucose levels, thereby restoring the pathogen-binding efficiency of MNVs. We measured blood glucose levels in the blood of diabetic rats after repetitive blood-cleansing with hRBC-MNVs (Figure 2k). The blood glucose levels gradually decreased from ∼380 to ∼280 mg dL^-1^ after three rounds of blood-cleansing, which recouped the MRSA depletion efficiency after the second round of blood-cleansing with hRBC-MNVs, where the blood glucose level was approximately 300 mg dL^-1^ (Figure 2l).

Finally, we validated the binding diversity of hRBC-MNVs and hWBC-MNVs using human fecal material and variants of SARS-CoV-2, respectively. The fecal material containing 168 bacterial species spiked in human whole blood mimics bacteremia triggered by fecal peritonitis. hRBC-MNVs magnetically depleted 99.97% of the polymicrobial fecal microbiome spiked in human whole blood, of which 135 species among the 168 bacterial species were effectively captured by hRBC-MNVs (Figure 2m). The magnetic removal efficiency of the bacterial species was greater than 90%: it was 100% in 50 species, 99.9– 99.99% in 72 species, 90–99.9% in 9 species, and 90–99% in 5 species (Table S1). Notably, the binding efficiencies of hWBC-MNVs to the spike proteins of SARS-CoV-2 were consistent regardless of their mutations, including B.1.1.7, B.1351, B.1.617.2 and B.1.1.529 (∼70%) (Figure 2n). This could be attributed to the conserved glycosylation sites of the spike protein in all variants opsonized by MBL, which were then effectively captured by CR1 and CR3 on the hWBC-MNVs.^[60]^

### 2.2 Magnetic blood-cleansing system

We employed the improved microfluidic magnetic separation system to demonstrate the depletion of a broad range of pathogens and proinflammatory cytokines in the blood using MNVs. The magnetic blood-cleansing system consists of two major components: a microfluidic blood-cleansing device for magnetically separating MNV-bound pathogens and cytokines and a series of flexible tubing knots to continuously mix the infused MNVs with blood.^[61]^ The single inlet channel branches three times before entering the eight magnetic separation channels (Figure S3). To enhance the magnetic separation of MNV-bound pathogens in viscous fluids, such as whole blood, an array of slanted obstacles inducing secondary spiral flows in the microfluidic channels^[62]^ was integrated into the microfluidic channels, and a Halbach array was placed underneath the device to generate augmented magnetic flux density gradients for an extended area (**Figure 3a**).^[63,64]^ We confirmed that MNVs in whole blood were completely removed by the magnets while they passed through the device at a flow rate of 10 mL h^-1^ (Figure S4), and the high-throughput blood-cleansing capability was validated even at a flow rate of 6000 mL h^-1^, which should be sufficient for treating human patients in the future (Figure S5).

**Figure 3.**
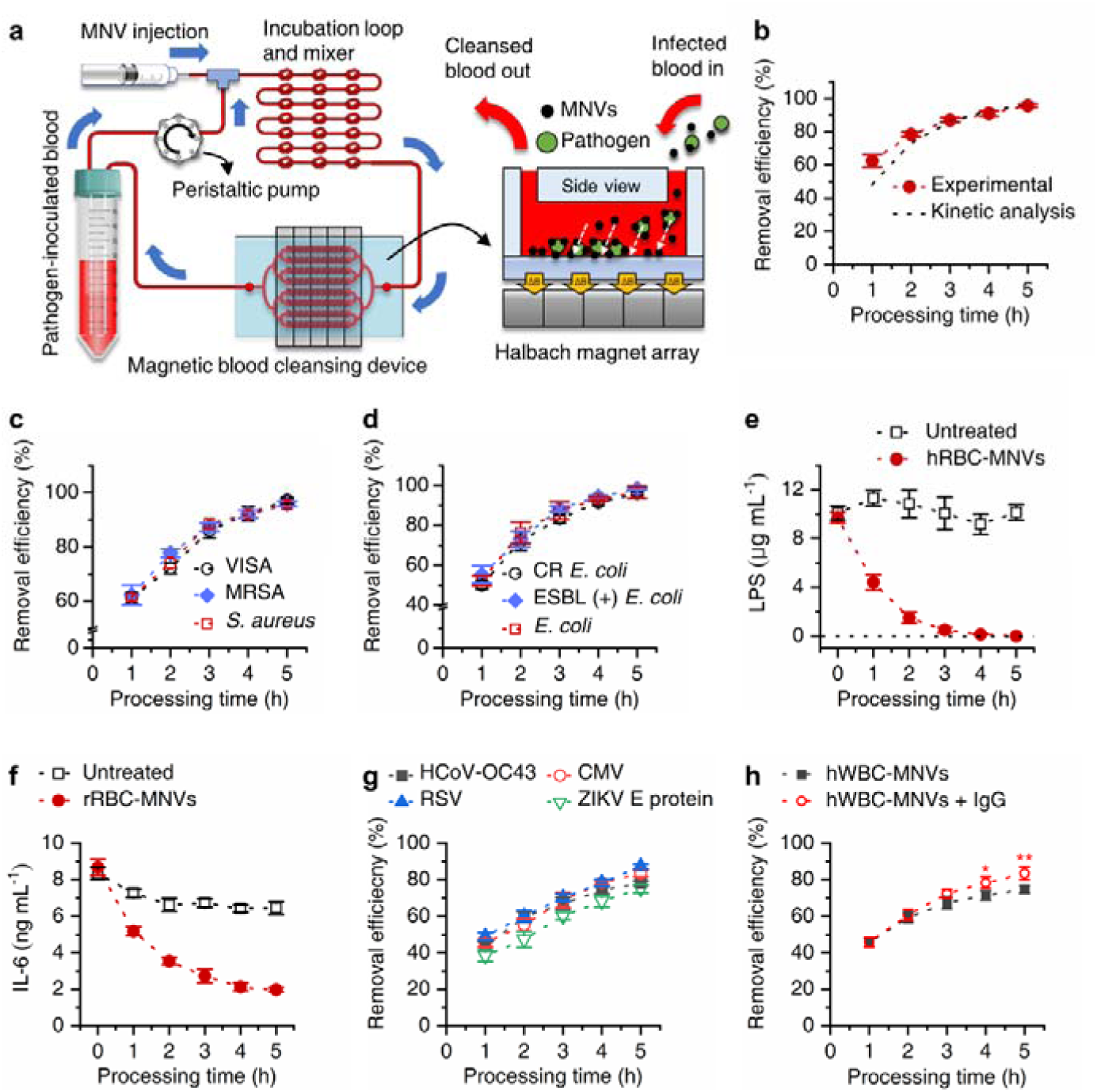
Continuous removal of pathogens and cytokines in the bacteremic blood using a blood-cleansing system in vitro. a) A blood-cleansing system for in vitro pathogen removal. Pathogen-inoculated blood mixed with MNVs flows into the incubation loop. After flowing through the mixing component, the pathogen-bound MNVs are magnetically captured on the bottom of the magnetic blood-cleansing device by the Halbach magnet array placed under the device. b) The removal efficiency of MRSA calculated by the Monod kinetics model corresponded to the experimental results for human blood (*n* = 3). c,d) The removal efficiency of antibiotic-resistant bacterial strains using hRBC-MNVs. The magnetic depletion rates of c) *S. aureus*, vancomysin-intermediate *S. aureus* (VISA), and MRSA and d) *E. coli*, ESBL-EC, and carbapenem-resistant *E. coli* (CR *E. coli*) using hRBC-MNVs in human blood. e,f) The removal efficiency of e) LPS using hRBC-MNVs and f) interleukin-6 (IL-6) using rRBC-MNVs. g) Removal efficiency of HCoV-OC43, CMV, RSV, and Zika virus (ZIKV) E protein using hWBC-MNVs in human blood. h) Removal efficiency of SARS-CoV-2 S protein using hWBC-MNVs was augmented by supplementing anti-SARS-CoV-2 S protein IgG in human blood. Data were expressed as means ± SEM. \**P* < 0.05; \*\**P* < 0.005 (a two-tailed Student’s t test).

For simulating extracorporeal treatments of animals infected with pathogens in vitro, we first circulated 10 mL of the blood spiked with pathogens through the blood-cleansing system at a flow rate of 10 mL h^-1^ while perfusing MNVs into the line (Figure 3a). Approximately 79% of MRSA in the blood was removed every hour, and over 95% was eradicated within 5 hours, corresponding to the theoretical prediction based on the Monod kinetics model (Figure 3b). We also tested whether the acquisition of antibiotic resistance in gram-positive (*S. aureus*, MRSA, and vancomycin-intermediate *Staphylococcus aureus* (VISA)) and gram-negative (*E. coli*, ESBL-positive *E. coli*, and carbapenem-resistant (CR) *E. coli*) bacteria compromises the capturing efficiency of MNVs due to alteration of the recognized molecules present on the bacterial cell wall. Despite the reported modification of PBP2a on MRSA and cell wall peptidoglycan on VISA,^[65,66]^ which was induced by the acquisition of antibiotic resistance, the capturing efficiency of hRBC-MNVs to all three strains was retained (Figure 3c), which could be attributed to the interaction between the glycophorins and the mucin-binding proteins in *S. aureus*.^[67-69]^ Similar binding characteristics were also observed for the gram-negative bacteria in which *E. coli*, ESBL-positive *E. coli*, and CR *E. coli* were depleted in the blood at the same rate (Figure 3d). This was because glycophorins on hRBC-MNVs bind with fimbrial adhesins other than the outer membrane porins known to be modified upon the acquisition of antibiotic resistance in *E. coli*.^[70-72]^

The removal efficiency of lipopolysaccharide (LPS) in the blood was even more proficient than that of intact bacteria, and LPS was completely eradicated within 5 hours (Figure 3e) because the diffusion coefficient of LPS is approximately 10^4^ times larger than that of bacteria due to their smaller size^[73,74]^; thus, the capturing efficiency is increasing.^[23]^ Moreover, we demonstrated that DARC on RBC-MNVs depleted interleukin-6 (IL-6) in whole blood for 5 hours (Figure 3f), and IL-6 is one of the proinflammatory cytokines strongly associated with the pathogenesis of sepsis.^[75,76]^

As we validated the virus depletion capability of hWBC-MNVs in Figure 2i, the blood samples (10 mL) spiked with three viruses (HCoV-OC43, CMV, and RSV) and two viral envelope proteins (Zika virus E protein and SARS-CoV-2 S protein) were continuously treated by the blood-cleansing device, and 70-80% of the viruses or the viral proteins were removed within 5 hours (Figure 3g,h). Moreover, when blood containing the SARS-CoV-2 S protein was supplemented with anti-SARS-CoV-2 S protein antibody, we obtained improved depletion efficiency after 5 hours of treatment (*P*< 0.05) (Figure 3h) because the Fc receptors, in addition to CR1 and CR3 on hWBC-MNVs, provided adjuvant capturing capability via SARS-CoV-2 S protein-specific antibodies.

### 2.3 Extracorporeal blood-cleansing treatment of bacteremia models

To validate the potential clinical impact of our MNV-based blood-cleansing approach, which improves mortality while restoring immune homeostasis, we used rat bacteremia models induced by MRSA and CR *E. coli* infection. The cleansing device was linked to the jugular veins of anesthetized rats via catheters (**Figure 4**a). We did not include the experimental condition for the control group, in which the infected rats were treated with null MNPs, because a previous study already demonstrated that MNPs without any binding capacity do not confer any therapeutic effects (10). The MRSA-infected rats receiving the 5-hour blood-cleansing treatment using rat RBC (rRBC)-MNVs had MRSA levels that gradually decreased in the blood (99% depletion), while the untreated animals had consistently high levels of MRSA (> 10^5^ CFU mL^-1^ in blood) (Figure 4b). However, bacterial concentrations in the blood rebounded within the next 24 hours, presumably due to their growth in the blood and translocation from organs to the bloodstream. We provided those rats with another 5-hour blood-cleansing treatment on the following day, and MRSA levels in the blood also gradually diminished, reaching an undetectable level at the fifth hour of treatment (Figure 4b). The peripheral capillary oxygen saturation of the treated rats (> 95%) was distinguishable from that of the untreated animals (< 90%) (Figure 4c), which could be associated with organ dysfunction due to hypoxemia.^[77]^ The proinflammatory cytokine levels in the blood, including TNF-α, IL-6, IL-1β, GM-CSF, and IL-4, significantly decreased even after a single round of 5-hour treatment, which could have resulted from the depleted bacteria in the blood and were directly removed by DARC on rRBC-MNVs (Figure 4d). All untreated rats died within 16 hours, and those treated with one 5-hour treatment died within two days. However, the rats that received two serial 5-hour treatment rounds for two days fully recovered within seven days, and those that survived for seven days were humanely euthanized according to the IACUC protocol (Figure 4e). The WBC counts and the platelet levels in the blood completely recovered seven days postinfection once the rats received the two-serial blood-cleansing treatments for two days (Figure 4f,g). Most importantly, immunohistochemical analysis showed that two serial rounds of 5-hour blood-cleansing significantly reduced the pathogen loads in the lungs, spleens, and kidneys of rats (Figure 4h,i). These pathogen reductions in the major blood clearance organs could account for the prominent reduction in the mortality of rats treated with blood-cleansing (Figure 4e).

**Figure 4.**
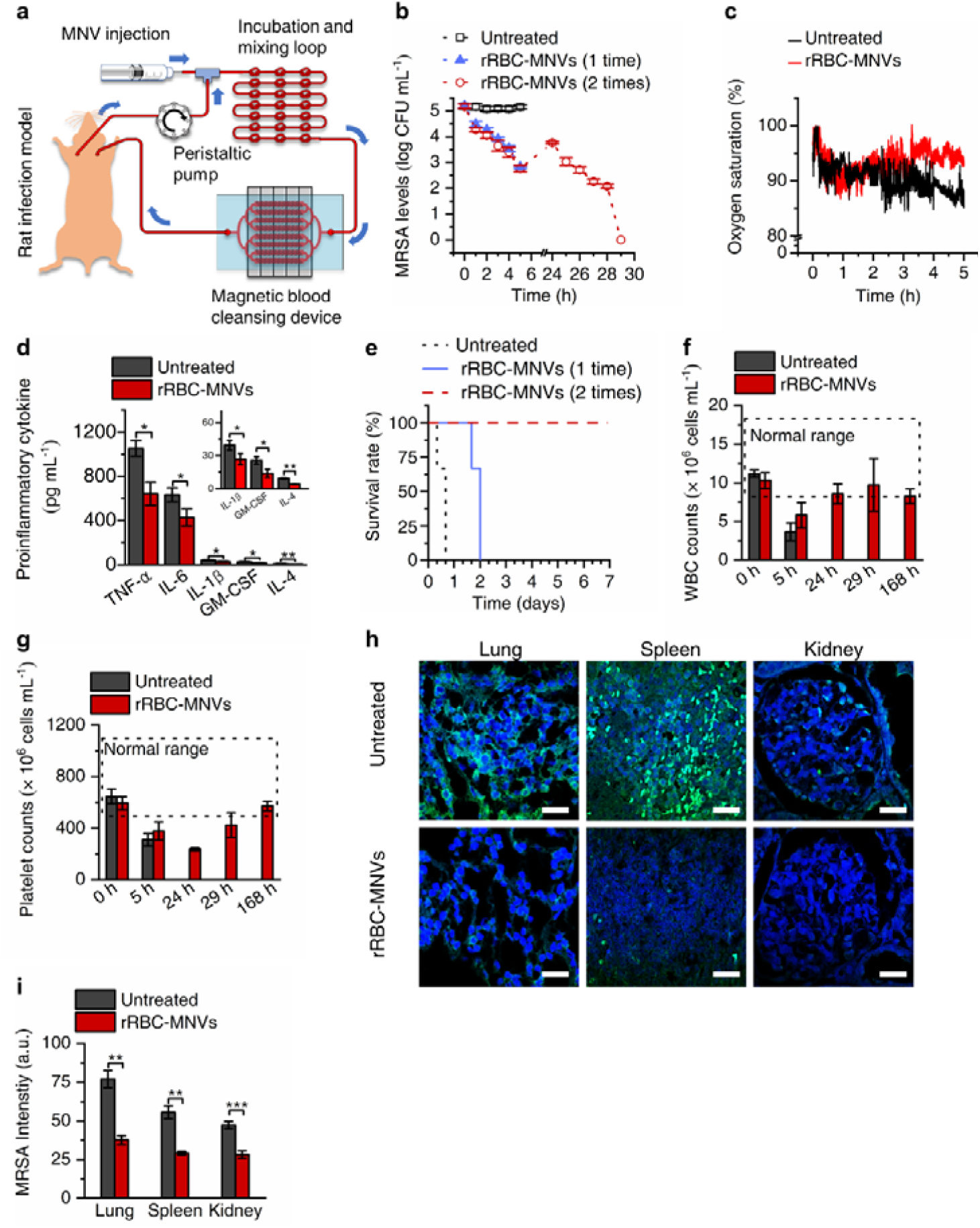
Extracorporeal magnetic blood-cleansing treatment in MRSA-infected rats and consequent restoration of immune homeostasis. a) A Scheme of an experimental setup for extracorporeally removing pathogens and proinflammatory cytokines in lethally infected bacteremic rats using RBC-MNVs. b) The bacterial levels in the MRSA-infected rats over the time course of the 5-hour extracorporeal treatment (*n* = 3) in comparison to that of the untreated group (*n* = 3). c) The oxygen saturation levels in the MRSA-infected rats during the rRBC-MNV-based blood-cleansing treatment for 5 hours. d) The proinflammatory cytokine levels in the MRSA-infected rats that received the rRBC-MNV blood-cleansing treatment (*n* = 3) significantly decreased compared to that of the control group (*n* = 3). e) Kaplan-Meier plots represent the increased survival of the MRSA-infected rats when they were treated by a single round of 5-hour blood-cleansing (solid blue line, *n* = 3) and two serial rounds of 5-hour treatment for 2 days (dashed red line, *n* = 3) compared to that of the untreated group (black dotted line, *n* = 3). f) The WBC counts and g) platelet counts of the MRSA-infected rat models that received 5-hour extracorporeal blood-cleansing using rRBC-MNVs. Both blood components represent a restoration of immune homeostasis that was recovered after receiving two serial 5-hour blood-cleansing treatments for 2 days. h) Immunofluorescence images of the lung, spleen, and kidney tissue slices obtained from MRSA-infected rats that were either untreated (top) or treated (bottom) by two serial rounds of 5-hour blood-cleansing for 2 days. The samples were stained with anti-MRSA antibody (green) and Hoechst (blue). Scale bar, 20 μm. i) MRSA levels in the major organs were quantified from the images obtained from six random fields. The MRSA levels in the major organs receiving the rRBC-MNV blood-cleansing treatment (*n* = 3) were significantly reduced compared to those in the untreated group (*n* = 3). Data were expressed as means ± SEM. Statistical significance was calculated by a two-tailed Student’s t test. \**P* < 0.05; \*\**P* < 0.005; \*\*\**P* < 0.001.

We then administered a lethal dose of CR *E. coli* to the rats to validate the superiority of our system over antibiotic treatment. The untreated rats consistently had CR *E. coli* levels over 10^5^ CFU mL^-1^ in their blood for the first five hours, whereas intravenous administration of colistin (1 mg kg^-1^) effectively decreased the bacterial levels for five hours. The depletion of the blood-cleansing device was more efficient in removing CR *E. coli* than MRSA (Figure 4b and **Figure 5**A) because MBL in the blood more effectively binds *E. coli* than *S. aureus*,^[78]^ thus causing rRBC-MNVs to more prominently capture CR *E. coli*. On the following day, the bacterial blood concentration increased to ∼10^4^ CFU mL^-1^; however, the second round of the 5-hour treatment completely eradicated CR *E. coli* in the blood. Although colistin treatment ameliorated the bacterial levels (Figure 5a), it exacerbated the endotoxin levels in the blood due to the bactericidal effects of antibiotics that produce a large amount of fragmented gram-negative bacteria, which in turn triggers endotoxemia (Figure 5b).^[79]^ However, because the blood-cleansing treatment using rRBC-MNVs physically eradicated both intact antibiotic-resistant bacteria and their bacterial cell wall debris, the treated rats persistently retained low endotoxin levels during the extracorporeal treatment. The colistin administration also induce a proficient increase in the blood proinflammatory cytokine levels, which are even higher than that of the rats without any treatment (Figure 5c). However, because treatment with rRBC-MNVs enables the simultaneous elimination of both pathogens and proinflammatory cytokines, the rats showed a significant reduction in major proinflammatory cytokines even after the first 5-hour treatment with rRBC-MNVs (Figure 5c). Two rounds of the 5-hour treatment for two serial days resulted in full recoveries from the severe infection within seven days; in contrast, the rats without blood-cleansing died within three days even with antibiotic treatment (Figure 5d). The treatment also enabled the WBC counts and platelet levels to return to the normal range within seven days postinfection (Figure 5e,f). The decreased WBC counts primarily associated with lymphocytopenia (Figure S6) could have been responsible for the high mortality of the rats that died within 2∼3 days.^[80,81]^ The recovery of the platelet levels in both MRSA- and CR *E. coli*-infected rats took more than 29 hours (Figure 4g, Figure 5f) because platelets are involved not only in acute response to infection but also in repairing damaged tissues and organs for recovery,^[82,83]^ which takes longer than remission of the WBC levels. Similar to the MRSA-infected rats receiving the blood-cleansing treatment (Figure 4h), the bacterial loads in the lungs, spleens, and kidneys of the CR *E. coli*-infected rats were significantly reduced compared to those of the other rats (Figure 5g,h). Most importantly, we analyzed the transcriptomic response of leukocytes in rats infected with CR *E. coli* to examine whether blood treatment using rRBC-MNVs could restore immune homeostasis. Among the differentially expressed genes (DEGs), which were generated by comparing the gene expression of CR *E. coli*-infected rats that received two serial rounds of 5-hour blood-cleansing to that of the untreated rats (Table S2), we selected DEGs with a log2-fold change value greater than 1.5 and a *P* value of less than 0.05 (Table S3). After the selection, sepsis-associated genes were collected from the DEGs using the Open Targets Platform (Figure 5i, Table S4). Notably, 83 genes among the 420 significantly downregulated genes were associated with sepsis, such as S100a8, -9,^[84]^ Lcn2,^[85]^ Pf4,^[86]^ Gzma, Gzmb,^[87]^ and Olfm4,^[88]^ which are known to be upregulated in septic or severe trauma patients; in contrast, only one gene out of the 10 significantly upregulated genes was identified as sepsis-associated by the Open Targets Platform, which was RT1-Da.^[89]^ Similar to RT1-Da, several other upregulated genes were found to be downregulated in septic patients, including CD74,^[90]^ Rps27, Rpl17, and Rpl3 (Figure 5i).^[91,92]^ Moreover, for the top 10 terms of gene ontology analysis in the downregulated biological process, differentially expressed genes with the highest significance in comparison to those in the untreated animals (*P* < 0.001) were associated with the response to external stimulus, response to stress, cell death and biological process involved in interspecies between organisms, supporting the potential clinical impact of our blood-cleansing system, which not only improves mortality but also reinstates the homeostasis of immunity (Figure 5j).

**Figure 5.**
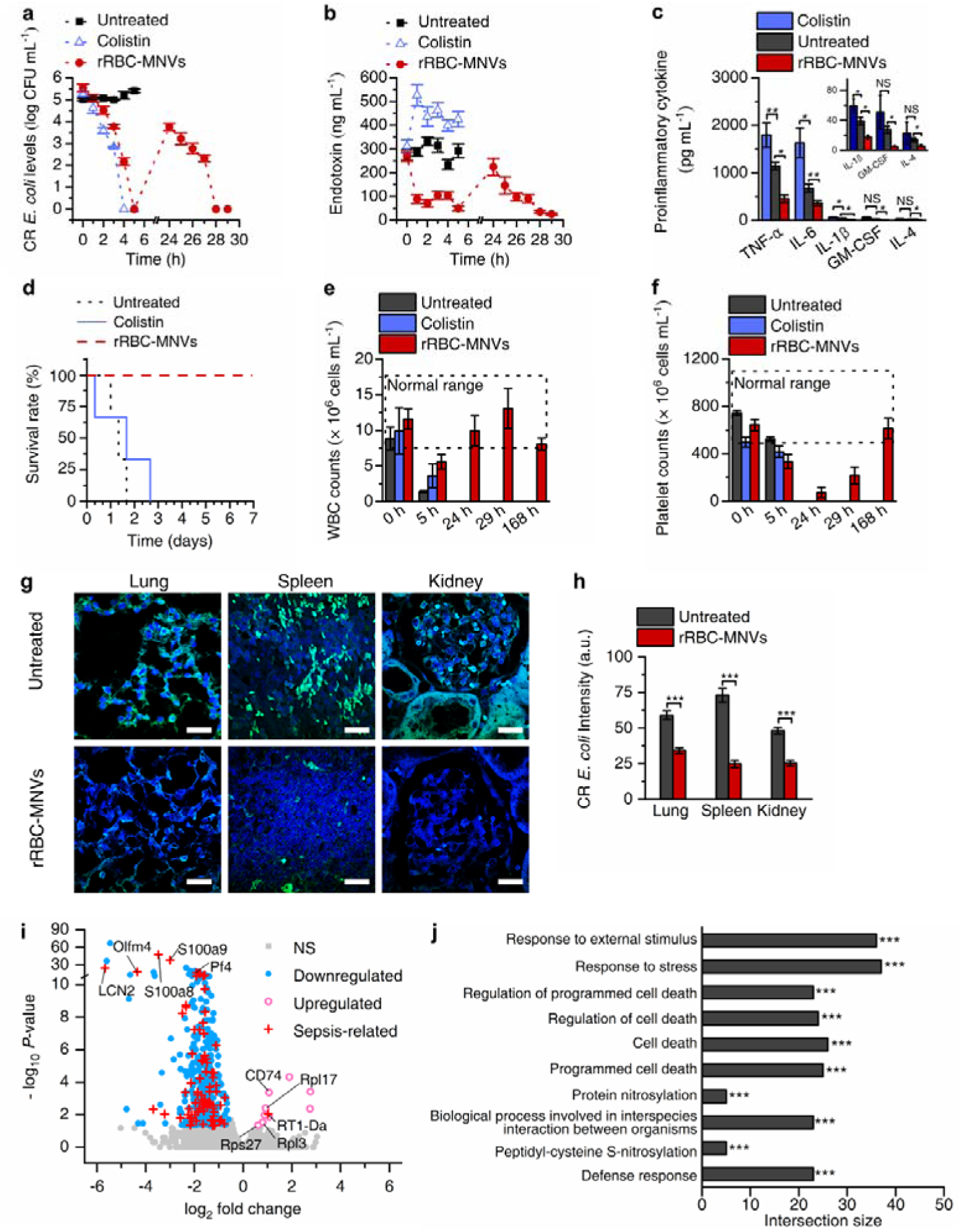
Extracorporeal magnetic blood-cleansing treatment in CR *E. coli*-infected rats and consequent restoration of immune homeostasis. a) The CR *E. coli* and b) endotoxin levels in the blood of the CR *E. coli*-infected rats, which were untreated, colistin-administered, and treated by two serial rounds of the 5-hour blood-cleansing treatment using rRBC-MNVs (*n* = 3 for each group). c) The proinflammatory cytokine levels in the blood of the CR *E. coli*-infected rats receiving no treatment (*n* = 3), colistin (*n* = 3), and blood-cleansing treatment using rRBC-MNVs (*n* = 3). d) Kaplan-Meier plots represent the increased survival in the CR *E. coli*-infected rats that received two serial rounds of the 5-hour blood-cleansing treatment using rRBC-MNVs (red dashed line, *n* = 3) compared to that of the untreated group (black dotted line, *n* = 3) and the colistin-administered group (solid blue line, *n* = 3). e) The WBC counts and f) platelet counts of the CR *E. coli*-infected rats with and without the two serial rounds of the 5-hour blood-cleansing treatment. g) Immunofluorescence images of the lung, spleen, and kidney tissue slices obtained from CR *E. coli*-infected rats that were untreated (top) or treated (bottom) by two serial rounds of 5-hour blood-cleansing for 2 days. The samples were stained with anti-*E. coli* antibody (green) and Hoechst (blue). Scale bar, 20 μm. h) CR *E. coli* levels in the major organs were quantified from the images obtained from six random fields. The CR *E. coli* levels in the major organs that received the rRBC-MNV blood-cleansing treatment (*n* = 3) were significantly reduced compared to those in the untreated group (*n* = 3). i) Volcano plot of the differentially expressed genes (DEGs) in the CR *E. coli*-infected rats that received two serial rounds of 5-hour blood-cleansing in comparison to those of the untreated rats. The significantly upregulated (pink empty circles) and downregulated DEGs (blue dots), including sepsis-associated genes, were distinguished from other DEGs (gray dots) (*P* < 0.05 & log_2_ fold change > 1.5). j) Gene ontology categories of the top ten enriched biological processes in the CR *E. coli*-infected rats receiving two serial rounds of the 5-hour blood-cleansing treatment support that the blood-cleansing treatment facilitates suppression of the biological processes associated with sepsis. Data were expressed as means ± SEM. Statistical significance was calculated by a two-tailed Student’s t test. \**P* < 0.05; \*\**P* < 0.005; \*\*\**P* < 0.001; NS, not significant.

## 3. Conclusion

A few extracorporeal blood-cleansing devices have been used in clinical settings as adjuvant therapeutic tools to treat infected patients; however, contentions about the therapeutic efficacy of these devices have occurred as their ability to simultaneously eradicate a broad spectrum of inflammation-triggering reagents, including bacteria, viruses, endotoxin, and cytokines, has been imperfect.^[22,93]^ Recent efforts have been made in conjunction with nanobiotechnology and microfluidics, and a new therapeutic strategy was proposed^[10,23,94]^; however, the strategy lacks the ability to remove clinically relevant pathogens prior to diagnosis because the binding targets permitted by a certain type of antibody or opsonins are inherently limited.^[24]^ We report for the first time that a significant reduction in both pathogenic materials and proinflammatory cytokines could lead to full recovery from severe infection, and this result was corroborated by survival rates, histological analysis, blood counts, and transcriptomic analysis.

Our approach provides an unparalleled level of removal efficiency and binding target ranges in human whole blood while avoiding the potential immune response caused by MNPs because the MNPs are camouflaged by blood cells, which have been demonstrated as immunologically inert.^[95]^ MNV multifunctionality was induced by a combination of the surface receptors on the corresponding blood cells, such as GYPA, DARC, CR1, and CR3, which facilitate the simultaneous removal of a wide range of multidrug-resistant bacteria, pandemic-potential viruses, endotoxin, and proinflammatory cytokines without requiring previous knowledge on their identities. This unique capability would be very useful when treating patients with superinfections because complications due to viral infection often accompany bacterial infections,^[96]^ contributing to the high mortality. Moreover, the depletion of unculturable bacteria in the blood is also crucial for treating patients with bloodstream infection because the conventional diagnostic method cannot detect the presence of those bacteria.^[12]^

The MNV depletion efficiently targets pathogens and can be further augmented since we unveiled the quantitative role of the receptors present on MNVs, which involves capturing pathogens, as well as their synergistic effects when combined with the use of opsonins or antipathogen immunoglobulins. For example, patients with low levels of native opsonin, such those inherently exhibiting low blood MBL levels (30% of the population worldwide),^[97]^ those with Behcet’s disease with low serum MBL,^[98]^ and COVID-19 patients with low C1q in the blood,^[99]^ could maximize their therapeutic efficiency using MNVs by infusing insufficient opsonin molecules (e.g., MBL or ficolin-1), the blood plasma of healthy donors or the blood plasma of those who are vaccinated against the pathogen (IgG) into the extracorporeal circuit, as these additions supplement the deficient binding capability of MNVs.

Moreover, we can reasonably extend our system to other infectious diseases, such as HIV and malaria infections, because HIV is known to interact with CD4 on T cells,^[43]^ DARC on RBCs,^[100]^ and blood MBL,^[101]^ and Plasmodium falciparum has been shown to bind GYPA^[102]^ and CR1,^[103]^ which implies that these pathogens could also be removed by either WBC-MNVs or RBC-MNVs. Finally, our system can provide a potential life-saving treatment method for COVID-19 patients by simultaneously reducing the SARS-CoV-2 virus and its variants, blood proinflammatory cytokines, because the deaths from COVID-19 are closely associated both with the viral loads in the blood^[17]^ and the excessive proinflammatory cytokines in the blood.^[104,105]^

## 4. Experimental Section

### Preparation of blood cells

To prepare RBCs, WBCs, and PLTs, human whole blood samples were obtained from the Republic of Korea National Red Cross (South Korea) (UNISTIRB-19-23-C), and rat whole blood samples were collected into a heparinized tube (BD Bioscience, San Jose, CA, USA) (UNISTIACUC-20-51). RBCs were harvested at 800 *g* for 5 min at 4 °C from whole blood samples and resuspended in 1× PBS. For the WBCs, the human blood samples were added to ACK lysis buffer at a ratio of 1:10 and incubated at room temperature for 5 min, and the mixtures were centrifuged at 300 *g* for 5 min at 4 °C. The supernatant was discarded, and the pellets were resuspended in a cold 1× PBS solution followed by centrifugation at 300 *g* for 5 min at 4 °C. The pelleted WBCs were resuspended into 1× PBS. PLTs were also isolated from human blood samples with two-step centrifugation. The whole blood samples were centrifuged at 100 *g* for 10 min. The top layer of plasma was transferred into a new tube following centrifugation at 400 *g* for 10 min, and the pelleted PLTs were resuspended in 1× PBS solution. The different types of isolated blood cells were used to prepare their membrane-derived MNVs within an hour to prevent the inactivation of the membrane proteins.

### Cell membrane derivation

To prepare RBC membrane ghosts, the RBCs were washed 3 times with 1× PBS buffer (pH 7.2, Biosesang, Seongnam, South Korea) followed by incubating the RBC pellets with a 25% v/v mixture of PBS and distilled water (Biosesang, Seongnam, South Korea) to treat the cell membrane hypotonically for 1 hour at 4 °C. The RBC membrane ghosts were purified by removing hemoglobin with centrifugation at 14,000 rpm for 5 min and resuspending in a cold 1× PBS solution. Next, WBCs, HL60 cells, U937 cells, and hHSECs were incubated for 1 hour in hypotonic lysis buffer containing 30 mM Tris-HCl and 225 mM D-mannitol, 75 mM sucrose, 0.2 mM ethylene glycol-bis(2-aminoethylether)-N,N,N′,N′-tetraacetic acid (EGTA) and protease inhibitor cocktails (all reagents from Sigma-Aldrich, MO, USA) in DI water. PLT membranes were prepared by a three-time repeated freeze-thaw process. PLTs in 1× PBS were frozen at - 80 °C for 15 min and then thawed at room temperature, which was followed by centrifugation at 800 *g* for 3 min and washing with 1× PBS. The hypotonically treated cell membranes were then ultrasonicated (22 kHz at 100 W for 5 min) to form the membrane-derived vesicles without intracellular organelles.

### Preparation of MNVs

One milligram of MNPs (Ademtech, Carboxyl-Adembeads, 200 nm in diameter, Pessac, France) and membrane vesicles derived from 10^5^ of each type of blood cells and cell lines were extruded sequentially through 1-μm, 0.4-μm, and 0.2-μm pore size track-etched membrane filters (Sterlitech, WA, USA) installed on an Avanti mini extruder (Avanti Polar Lipids, AL, USA). After the extrusion process, the MNVs were magnetically purified by removing the supernatant and were stored in 1× PBS at 4 °C for further studies.

### Characterization of MNVs

The hydrodynamic diameter and the surface charge of MNVs were measured through dynamic light scattering (DLS) (Nano ZS, Malvern analytical, Malvern, UK). The morphology of MNVs negatively stained with 1% phosphotungstic acid (Sigma-Aldrich, MO, USA) was visualized with transmission electron spectroscopy (JEM-1400, JEOL, Japan). To isolate the inherited membrane proteins from MNVs, we collected RBC-MNVs (or WBC-MNVs), which were synthesized from 1 mg MNPs and 10^5^ cells and resuspended in 0.5 mL RIPA buffer (Biosesang, Seongnam, South Korea) with a protease inhibitor tablet (Thermo Fisher Scientific Inc, MA, USA), which was followed by incubation for 30 min at 4 °C. The resulting solutions were centrifuged at 13,000 rpm for 10 min at 4 °C, and the supernatant (0.5 mL) containing cell membrane molecules was diluted into 0.5 mL cold 1× PBS. The molecules (GYPA, CR1, DARC, and CR3) extracted from MNVs, which capture a broad range of pathogens and proinflammatory cytokines, were quantified by using ELISA kits (GYPA: LS-F9587, LSBio, WA, USA; CR1: ab277439, Abcam, MA, USA; DARC: MBS450049, MyBioSource, CA, USA; CR3: ab277412, Abcam, MA, USA).

### Cell culture

U-937 cell line (21593.1, Korea Cell Line Bank, KCLB, Seoul, South Korea) was cultured in RPMI-1640 medium (Welgene, Gyeongsangbuk-do, South Korea) supplemented with 10% fetal bovine serum (Welgene, Gyeongsangbuk-do, South Korea) and 1% penicillin (Welgene, Gyeongsangbuk-do, South Korea). The HL-60 cell line (10240, Korea Cell Line Bank, KCLB, Seoul, South Korea) was cultured in DMEM (Welgene, Gyeongsangbuk-do, South Korea) supplemented with 20% fetal bovine serum and 1% penicillin. Human hepatic sinusoidal endothelial cells (PM10652, Innoprot, Vizcaya, Spain) were cultured in endothelial cell medium (Innoprot, Vizcaya, Spain) supplemented with 1% endothelial cell growth supplement (ECGS, Innoprot, Vizcaya, Spain), and penicillin-streptomycin solution (Innoprot, Vizcaya, Spain). All cells were cultured in a cell incubator at 37°C with 5% CO_2_, and each cell culture medium was refreshed every one or two days, depending on the cell growth. To differentiate U937 cells into M0 macrophage-like cells, 100 ng mL^-1^ phorbol-12-myristate-13-acetate (PMA, Sigma-Aldrich, MO. USA) was added to U937. After two days, the fresh culture media were replaced, and the differentiation was completed on the following day. To differentiate HL60 cells into neutrophil-like cells, 0.1 nM all-trans retinoic acid (ATRA, Sigma-Aldrich, MO. USA) were added to HL60. After two days, the fresh culture medium was supplemented with 0.1 nM ATRA and the the differentiation was completed on the following day.

### Preparation of pathogens

Extended-spectrum β-lactamase-positive *Escherichia coli* (ESBL-positive *E. coli*) (CCARM 1341) and methicillin-resistant *Staphylococcus aureus* (MRSA) (CCARM 3140) were purchased from CCARM (Culture Collection of Antimicrobial Resistant Microbes, Gyeonggi-do, South Korea). *Staphylococcus aureus* (*S. aureus*) was purchased from KCCM (Korean Culture Center of Microorganisms, Seoul, South Korea). Carbapenem-resistant *E. coli* (CR *E. coli*) and vancomycin-intermediate *S. aureus* (VISA) were obtained from the National Culture Collection for Pathogens (NCCP, Cheongju, South Korea). All bacteria were cultured on LB broth agar (LB) overnight at 37 °C. A single colony of each bacterial strain was incubated in LB broth medium at 37 °C and shaken 250 rpm until a stationary phase was reached. Prior to use in all experiments, the bacterial suspension was inoculated into fresh LB broth media and grown until an early exponential phase in the growth curve was reached. Human coronavirus 229E (KBPV-VR-9), human coronavirus OC43 (KBPV-VR-8), and human respiratory syncytial virus A (KBPV-VR) were obtained from the South Korean Bank of Pathogenic Viruses (KBPV, South Korea). Human coronavirus NL63 (NCCP 43214) was obtained from NCCP, South Korea. The viruses were stored in 100-μL aliquots at - 80 °C for further study. Lipopolysaccharide (LPS, *E. coli* serotype O111:B4)-endotoxin was purchased from Sigma-Aldrich (L3012, MO, USA). S proteins of SARS-CoV-2 (40589-V08B1, USA-WA1/2020) and its variants (40589-V08B6 B.1.1.7, 40589-V08B9 B.1351, 40589-V08B16 B.1617.2, 40589-V08H26 B.1.1.529) were purchased from Sinobiological (Beijing, China). The human fecal microbiome was purchased from BioBankHealing (Seongnam, South Korea).

### Measurement of the pathogen binding efficiency

The pathogen-binding efficiencies of MNVs derived from different cells were first evaluated in a test tube environment to select MNVs with the highest affinity to pathogens. The 10^4^ CFU of bacteria (or 10^4^ PFU of viruses) were spiked into 1 mL of human whole blood that was obtained from the Republic of Korea National Red Cross as described above. Then, the blood samples were transferred into a new 1.5 mL tube containing 0.15 mg of MNVs (or a new empty tube without MNVs as a control) and mixed for 15 min with steady agitation using an inverting mixer at room temperature. The blood samples were located next to the Halbach array (59) of N52-grade magnets (KJ Magnetics, PA, USA) for 10 min, and the supernatant of the blood sample was collected to measure the pathogen-binding efficiency. The pathogen-binding efficiencies (*I*_eff_) of MNVs were assessed based on the following equation where *C*_p_^MNV^ and *C*_p_^ctrl^ are the concentrations of MNVs and pathogens in the blood samples, respectively, and the procedure was performed as described above except there was no MNVs.

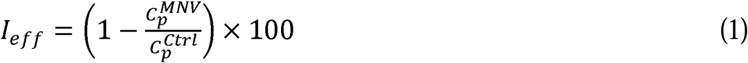

To measuring the quantitative role of GYPA, CR1, and CR3 in capturing each type of pathogen, GYPA and CR1 on RBC-MNVs and CR1 and CR3 on WBC-MNVs were individually or simultaneously blocked with their corresponding antibodies (GYPA: ab129024; CR1: ab133293; CR3: ab52920 and ab184308, Abcam, Cambridge, UK) by incubating them for 1 hour at room temperature. After incubation, the MNVs treated with each blocking antibody were magnetically washed with 1× PBS prior to measuring the pathogen binding efficiency in the blood.

### Measurement of the D-glucose-removal efficiency and pathogen-removal efficiency in hyperglycemic blood

Hyperglycemia was induced in 8-week-old Wistar rats (Orient Bio Inc, Seongnam, South Korea) by intraperitoneal (IP) injection of a single dose of streptozotocin (STZ) (65 mg kg^-1^, Sigma-Aldrich, MO, USA) after an IACUC approval (UNISTIACUC-18-03) was received. Then, the rats were housed and fed 10% sucrose water (S1888, Sigma-Aldrich, MO, USA) for 2 days. One week after the IP injection of STZ, blood samples were collected from the rats, and 8-hour fasting blood glucose levels were measured with a blood glucose meter (ACCU-CHEK Active, Roche, Basel, Switzerland). We confirmed that these rats had hyperglycemia as their blood D-glucose levels were greater than 125 dg mL^-1^. After the blood sample were collected, they were immediately stored at 4 °C and used for 2 hour. A total of 10^4^ CFU of MRSA was spiked into 1 mL of the hyperglycemic blood samples and the samples were incubated for 10 min. Then, RBC-MNVs (0.15 mg mL^-1^) were mixed with the blood sample, which was followed by three sequential magnetic separations in which we measured the glucose levels and MRSA concentrations in the supernatant after each magnetic separation. The blood glucose levels were measured with a blood glucose meter, and MRSA concentrations were quantified by plating 100 µL of the supernatant on LB agar plates.

### Fabrication of the magnetic blood-cleansing system

The designed magnetic separation devices were fabricated using a hot embossing process (LabEcon 300, Fontijne presses, Viaardingen, Netherlands) with epoxy stamps. In the first step, the designed channel patterns were positively micromachined on a PMMA substrate using a computer numerical control (CNC) milling machine (David 3020C, David, Incheon, South Korea). A mixture of PDMS precursor and the curing agent at a mass ratio of 10:1 was mixed and cast onto the PMMA micromachined template. After degassing the PDMS mixture to remove bubbles and curing for 2 hours at 60 °C, the PDMS molds were peeled off of the PMMA template. To fabricate the epoxy stamp, epoxy resin (Conapoxy FR-1080, Cytec Industries Inc, NY, USA) was poured into the PDMS mold and cured for 16 hours at 120 °C, which was followed by postcuring for 2 hours at 180 °C. After being cooled at room temperature, the epoxy stamp was demolded from the PDMS mold. For the hot embossing processes, a planar PMMA substrate was placed on the fabricated epoxy stamps and then pressed with 5 kN at 100 °C for 20 min, which was followed by cooling until 65 °C was reached. The hot-embossed PMMA channel substrates were then treated with isopropyl alcohol (99%, Samchun Pure Chemical, Gyeonggi-do, South Korea) and bonded with a thin PMMA film (250 µm in thickness) using a hot embossing machine.

The microscale mixing component for continuously incubating the blood with the injected MNVs consisted of serial knots (20 knots) of flexible Tygon® tubing (ID 0.508 mm, AAD04103, Saint Gobain PPL Corp., Paris, France) as described in a previous report (57). Then, the blood-cleansing systems were prepared by connecting the outlet of mixer units and the inlet of magnetic separation devices.

### Operating the extracorporeal system with MNVs in vitro

We quantitated how efficiently the extracorporeal system could remove MNVs in the whole blood while the MNVs were traveling through the magnetic separation channel area with the Habach magnets placed underneath. Ten milliliters of the blood sample was circulated through the blood-cleansing device circuit at a flow rate of 10 mL h^-1^ using a peristaltic pump (Reglo ICC, ISMATEC, Wertheim, Germany), and hRBC-MNVs were continuously injected into the circuit using a syringe pump (Fusion 200, Chemyx Inc, TX, USA) at a flow rate of 0.5 mL h^-1^. Twenty microliters of a blood sample was collected every hour, and the residual MNVs were measured by TGA (Thermogravimetric analyzer, Q500, TA instruments, DE, USA). Briefly, 20 µL of blood samples was placed on a TGA Pt sample pan and the temperature was raised from 25 to 650 °C. We quantitated the residual MNP amounts by measuring the mass remaining in the pan after 650 °C was reached because the rest of the blood components except for MNPs should have been thermally oxidized.

Next, we assessed the pathogen-removal efficiency of MNVs using an extracorporeal blood-cleansing device in vitro. Pathogens (10^4^ CFU mL^-1^ of bacteria and 10^4^ PFU mL^-1^ of viruses) were spiked into 10 mL of human whole blood samples anticoagulated with heparin (16 U mL^-1^) in a 50 mL conical tube. The blood samples containing the pathogens were immediately introduced into the blood-cleansing device circuit in vitro at a flow rate of 10 mL h^-1^ using the peristaltic pump, and 0.5 mg mL^-1^ MNVs were continuously injected into the extracorporeal circuit using the syringe pump at a flow rate of 0.5 mL h^-1^. The pathogens in the blood were captured by MNVs while they flowed through a mixing component and they were magnetically depleted when they passed through the microfluidic magnetic separation device. The cleansed blood samples returned to the blood reservoir and recirculated through the circuit for 5 hours. During the 5-hour blood-cleansing process, blood samples (100 µL) were collected from the outlet of the magnetic separation device to measure pathogen concentrations in the blood by counting CFU or quantifying viral rRNA loads using qPCR.

To measure the LPS depletion efficiency, 100 μg of LPS was spiked into 10 mL of human whole blood samples, and the blood samples flowed through the blood-cleansing system in vitro as described above. The concentrations of LPS were measured by an ELISA kit (LS-F55757-1, LSBio, WA, USA).

### Proinflammatory cytokine depletion tests

We obtained blood samples containing proinflammatory cytokines from the rats with cytokine release syndrome (CRS) that were challenged with LPS (5 mg kg^-1^, intravenous injection). The rat blood samples containing proinflammatory cytokines were continuously treated with the blood-cleansing device using RBC-MNVs for 5 hours, and IL-6 levels were measured by an ELISA kit (ab234570, Abcam, Cambridge, UK).

### Viral protein removal efficiency

The SARS-CoV-2 Spike protein (10 µg, 40589-V08B1, Sinobiological, Beijing, China) and Zika virus envelope protein (10 µg, 40543-V02H, Sinobiological, Beijing, China) were spiked into 10 mL of human whole blood, and the blood samples were treated with a blood-cleansing device at a flow rate of 10 mL h^-1^. Each viral envelope protein concentration was measured by a COVID-19 Spike Protein ELISA kit (ab274342, Abcam, MA, USA) and Zika virus SPH2015 envelope protein ELISA kit (KIT40543, Sinobiological, Beijing, China), respectively, using a microplate reader (Synergy Neo2, Bio Tek Instruments, VT, USA).

### Bacteremia models in rats

All animal experiments were approved by the Institutional Animal Care and Use Committee (IACUC) of Ulsan National Institute of Science and Technology (UNIST) (approval number: UNISTIACUC-20-51). Eight-week-old male Wistar rats (Orient Bio Inc, Seongnam, South Korea) were anesthetized by inhalation of 4% isoflurane (Kyongbo Pharm, Seoul, South Korea) and catheterized using rat 3 F jugular vein catheters (SAI-Infusion, IL, USA). Heart rates, SpO2, rectal temperatures, and respiratory rates were monitored using a physiological monitoring system (Cat. No., Harvard apparatus, MA, USA) during all bacteremia experiments. To develop a MRSA bacteremia model in rats, a bolus of 1 mL 0.9% normal saline (HK inno.N, Seoul, South Korea) containing 1 × 10^10^ CFU of MRSA was intravenously infused through the jugular vein catheter. To establish a CR *E. coli* bacteremia model, 5 × 10^9^ CR *E. coli* in 2.5 mL normal saline were continuously injected through the jugular vein catheter for 5 hours to maintain consistent bacterial levels in the blood.

### Extracorporeal blood-cleansing treatment of rats infected with antibiotic-resistant bacteria

Eight-week-old male Wistar rats were used to validate the therapeutic efficacy of our system in vivo. The inlet of extracorporeal blood-cleansing circuits was connected to one of the jugular vein catheters using a 21 G blunt needle and then to a Luer-Barb fitting connector (HV-95714-36, Materflex, IL, USA). The circuit outlet was connected to the other jugular vein catheter using the same tubing connection method. The blood was pumped through the extracorporeal circuit by the peristaltic pump at a flow rate of 10 mL h^-1^, and the 0.5 mg mL^-1^ RBC-MNVs in heparinized saline (32 U mL^-1^) was continuously introduced into the circuit by the syringe pump at a flow rate of 0.5 mL h^-1^ as described in the in vitro experimental setup. We collected 30 µL of the blood sample every hour to measure bacterial concentrations. One milliliter of blood was additionally obtained at the time points of 0 hour and 5 hours to analyze the complete blood count (CBC) and cytokine levels. It was not possible to assess CBC and cytokines each hour because drawing 1 mL of blood per hour from rats with severe infection was not permissible according to IACUC. We compensated for the volume of blood drawn from the rats by administering the same volume of saline solution. After the first round of 5-hour blood-cleansing treatment was completed, the rats were returned to cages where they were closely monitored and supported with analgesics according to the IACUC protocol. Following 24 hours, the rats were anesthetized by inhalation of 1-2% isoflurane, and the second 5-hour blood-cleansing treatment was performed. The rats untreated and treated with the blood-cleansing devices were closely monitored, and on the seventh day after the treatment, blood samples were collected to analyze CBC and cytokine levels, and the rats were humanely euthanized according to the IACUC protocol.

Bacterial loads in the blood were quantified by culture on agar plates, and CBC results were obtained from a hematology analyzer (VetScan HM5, ABAXIS, Zoetis, UK). To analyze cytokine levels, the collected blood sample was centrifuged at 500 *g* for 15 min to obtain plasma samples in which the cytokine levels were analyzed by Bioplex Pro Rat cytokine plex assay (Bio-Rad, CA, USA). The LPS concentrations of the blood collected from the CR-*E. coli* bacteremia model were assessed by an ELISA kit (LS-F55757-1, LSBio, WA, USA).

Transcriptomics analysis was performed by Macrogen Inc. (Seoul, South Korea). Briefly, the blood samples drawn from rats with and without blood-cleansing treatments were immediately collected, placed into sterile heparinized blood collection tubes (BD Biosciences, CA, USA) and stored at 4 °C before analysis. Total mRNA from WBCs was isolated with a QIamp RNA blood kit (Qiagen, Hilden, Germany) and treated with DNase to remove residual DNA. cDNA libraries were synthesized from the purified RNA using an Illumina TruSeq RNA library kit (Illumina Inc., CA, USA) and sequenced by an Illumina HiSeq 2000 (Illumina Inc., CA, USA) to generate raw reads. The sequenced raw reads were mapped to the rat reference genome and the differentially expressed genes (DEGs) between the untreated and blood cleansing treatment using rRBC-MNV groups were selected by the DESeq2 nbinomWaldTest. The results were analyzed to generate a volcano plot of DEGs and gene ontology term analysis.

### Immunohistochemistry analysis

At the endpoints of each animal experiment, the rats were humanely euthanized according to the IACUC protocol. The lungs, spleens and kidneys were collected, fixed in 4% paraformaldehyde solution (Biosesang, Seongnam, South Korea) and embedded in paraffin. Then, the embedded organ samples were sectioned with a microtome (RM225, Leica Microsystems, Wetzlar, Germany) and processed for immunohistochemistry (IHC). A rabbit anti-*E. coli* (ab137967, Abcam, Cambridge, UK) or a rabbit anti-*S. aureus* (ab20920, Abcam, Cambridge, UK) antibodies with fluorochrome (Alexa 488) conjugated anti-rabbit IgG (ab150077, Abcam, Cambridge, UK) were used to stain against each pathogen. All processed tissue samples were also stained with Hoechst 33342 (Thermo Fisher Scientific Inc, MA, USA). Fluorescence images of the tissue samples were examined using a confocal microscope (LSM780, ZEISS, Oberkochen, Germany). The residual pathogen levels in the organs were quantified using ImageJ (NIH, MD, USA) by measuring the green fluorescence intensity in six random spots in the tissue samples.

## Data Availability

All data produced in the present work are contained in the manuscript.

## Supporting Information

Supporting Information is available from the Wiley Online Library or from the author.

## Acknowledgements

S.J.P. and S.K. contributed equally to this work. This work was supported by the Samsung Research Funding and Incubation Center for Future Technology (SRFC-IT1602-02). JHK was supported by a UNIST research fund (1.220023.01).

## Author contributions

S.J.P., S.K. and J.H.K. conceived the idea, and S.J.P., S.K. and J.H.K. performed all experiments and analysis the experimental result with the assistance of M.S.L., B.H.J. and A.E.G., and S.J.P., S.K. and J.H.K. wrote and edited the manuscript

## Data and materials availability

All data are available in the main text or the supplementary materials.

Received: ((will be filled in by the editorial staff))

Revised: ((will be filled in by the editorial staff))

Published online: ((will be filled in by the editorial staff))

## References

[1] J. A. Kempker, G. S. Martin, Lancet 2020, 395, 168.

[2] M. Klompas, T. Calandra, M. Singer, JAMA 2018, 320, 1433.

[3] E. De Clercq, J. Clin. Virol. 2004, 30, 115.

[4] J. C. Garcia Casallas, H. Robayo-Amortegui, Z. Corredor-Rozo, A.M. Carrasco-Márquez, J. Escobar-Perez, J. Med. Case. Rep. 2019, 13, 141.

[5] M. E. A. de Kraker, A. J. Stewardson, S. Harbarth, PLoS Med. 2016, 13, e1002184.

[6] D. van Duin, K. S. Kaye, E. A. Neuner, R. A. Bonomo, Diagn. Microbiol. Infect. Dis. 2013, 75, 115; P. G. Higgins, C. Dammhayn, M. Hackel, H. Seifert, J. Antimicrob. Chemother. 2010, 65, 233; J.-M. Rodríguez-Martínez, L. Poirel, P. Nordmann, Antimicrob. Agents Chemother. 2009, 53, 4783.

[7] A. Uzan-Yulzari, O. Turta, A. Belogolovski, O. Ziv, C. Kunz, S. Perschbacher, H. Neuman, E. Pasolli, A. Oz, H. Ben-Amram, H. Kumar, H. Ollila, A. Kaljonen, E. Isolauri, S. Salminen, H. Lagström, N. Segata, I. Sharon, Y. Louzoun, R. Ensenauer, S. Rautava, O. Koren, Nat. Commun. 2021, 12, 443.

[8] J. H. Kang, M. Super, C. W. Yung, R. M. Cooper, K. Domansky, A. R. Graveline, T. Mammoto, J. B. Berthet, H. Tobin, M. J. Cartwright, A. L. Watters, M. Rottman, A. Waterhouse, A. Mammoto, N. Gamini, M. J. Rodas, A. Kole, A. Jiang, T. M. Valentin, A. Diaz, K. Takahashi, D. E. Ingber, Nat. Med. 2014, 20, 1211.

[9] J. W. Fjalstad, E. Esaiassen, L. K. Juvet, J. N. van den Anker, C. Klingenberg, J. Antimicrob. Chemother. 2018, 73, 569.

[10] M. S. Lee, H. Hyun, I. Park, S. Kim, D.-H. Jang, S. Kim, J.-K. Im, H. Kim, J. H. Lee, T. Kwon, J. H. Kang, medRxiv 2021, 2021.02.19.21251962.

[11] H. Chung, J. H. Lee, Y. H. Jo, J. E. Hwang, J. Kim, Shock 2019, 51.

[12] J. C. Marshall, P. M. Walker, D. M. Foster, D. Harris, M. Ribeiro, J. Paice, A. D. Romaschin, A. N. Derzko, Crit. Care 2002, 6, 342.

[13] E. S. Pronker, T. C. Weenen, H. Commandeur, E. H. J. H. M. Claassen, A. D. M. E. Osterhaus, PLoS One 2013, 8, e57755.*** U. Theuretzbacher, Int. J. Antimicrob. Agents 2009, 34, 15.

[14] J. Fajnzylber, J. Regan, K. Coxen, H. Corry, C. Wong, A. Rosenthal, D. Worrall, F. Giguel, A. Piechocka-Trocha, C. Atyeo, S. Fischinger, A. Chan, K. T. Flaherty, K. Hall, M. Dougan, E. T. Ryan, E. Gillespie, R. Chishti, Y. Li, N. Jilg, D. Hanidziar, R. M. Baron, L. Baden, A. M. Tsibris, K. A. Armstrong, D. R. Kuritzkes, G. Alter, B. D. Walker, X. Yu, J. Z. Li, B. A. Abayneh, P. Allen, D. Antille, A. Balazs, J. Bals, M. Barbash, Y. Bartsch, J. Boucau, S. Boyce, J. Braley, K. Branch, K. Broderick, J. Carney, J. Chevalier, M. C. Choudhary, N. Chowdhury, T. Cordwell, G. Daley, S. Davidson, M. Desjardins, L. Donahue, D. Drew, K. Einkauf, S. Elizabeth, A. Elliman, B. Etemad, J. Fallon, L. Fedirko, K. Finn, J. Flannery, P. Forde, P. Garcia-Broncano, E. Gettings, D. Golan, K. Goodman, A. Griffin, S. Grimmel, K. Grinke, C. A. Hartana, M. Healy, H. Heller, D. Henault, G. Holland, C. Jiang, H. Jordan, P. Kaplonek, E. W. Karlson, M. Karpell, C. Kayitesi, E. C. Lam, V. LaValle, K. Lefteri, X. Lian, M. Lichterfeld, D. Lingwood, H. Liu, J. Liu, K. Lopez, Y. Lu, S. Luthern, N. L. Ly, M. MacGowan, K. Magispoc, J. Marchewka, B. Martino, R. McNamara, A. Michell, I. Millstrom, N. Miranda, C. Nambu, S. Nelson, M. Noone, L. Novack, C. O’Callaghan, C. Ommerborn, M. Osborn, L. C. Pacheco, N. Phan, S. Pillai, F. A. Porto, Y. Rassadkina, A. Reissis, F. Ruzicka, K. Seiger, K. Selleck, L. Sessa, A. Sharpe, C. Sharr, S. Shin, N. Singh, S. Slaughenhaupt, K. S. Sheppard, W. Sun, X. Sun, E. Suschana, O. Talabi, H. Ticheli, S. T. Weiss, V. Wilson, A. Zhu R. The Massachusetts Consortium for Pathogen, Nat. Commun. 2020, 11, 5493.

[15] M. G. Davies, P. O. Hagen, Br. J. Surg. 1997, 84, 920.

[16] J.-L. Vincent, S. M. Opal, J. C. Marshall, K. J. Tracey, Lancet 2013, 381, 774.

[17] C. Monard, T. Rimmelé, C. Ronco, Blood Purif. 2019, 2 H. Shoji, Ther Apher Dial 2003, 7, 108.

[18] J. H. Kang, Biochip J. 2020, 14, 63.

[19] J. H. Kang, E. Um, A. Diaz, H. Driscoll, M. J. Rodas, K. Domansky, A. L. Watters, M. Super, H. A. Stone, D. E. Ingber, Small 2015, 11, 5657.

[20] T. F. Didar, M. J. Cartwright, M. Rottman, A. R. Graveline, N. Gamini, A. L. Watters, D. C. Leslie, T. Mammoto, M. J. Rodas, J. H. Kang, A. Waterhouse, B. T. Seiler, P. Lombardo, E. I. Qendro, M. Super, D. E. Ingber, Biomaterials 2015, 67, 382.

[21] P. Angsantikul, S. Thamphiwatana, Q. Zhang, K. Spiekermann, J. Zhuang, R. H. Fang, W. Gao, M. Obonyo, L. Zhang, Adv. Ther. 2018, 1, 1800016.

[22] A. V. Kroll, Y. Jiang, J. Zhou, M. Holay, R. H. Fang, L. Zhang, Adv. Biosyst. 2019, 3, 1800219.

[23] E. Ben-Akiva, R. A. Meyer, H. Yu, J. T. Smith, D. M. Pardoll, J. J. Green, Sci. Adv. 2020, 6, eaay9035; Y. Chen, M. Chen, Y. Zhang, J. H. Lee, T. Escajadillo, H. Gong, R. H. Fang, W. Gao, V. Nizet, L. Zhang, Adv. Healthc. Mater. 2018, 7, 1701366.

[24] C.-M. J. Hu, R. H. Fang, K.-C. Wang, B. T. Luk, S. Thamphiwatana, D. Dehaini, P. Nguyen, P. Angsantikul, C. H. Wen, A. V. Kroll, C. Carpenter, M. Ramesh, V. Qu, S. H. Patel, J. Zhu, W. Shi, F. M. Hofman, T. C. Chen, W. Gao, K. Zhang, S. Chien, L. Zhang, Nature 2015, 526, 118.

[25] H. L. Anderson, I. E. Brodsky, N. S. Mangalmurti, J. Immunol. 2018, 201, 1343.

[26] S. Vandendriessche, S. Cambier, P. Proost, P. E. Marques, Front. Cell Dev. Biol. 2021, 9.

[27] N. Bose, A. S. H. Chan, F. Guerrero, C. M. Maristany, X. Qiu, R. M. Walsh, K. E. Ertelt, A. B. Jonas, K. B. Gorden, C. M. Dudney, L. R. Wurst, M. E. Danielson, N. Elmasry, A. S. Magee, M. L. Patchen, J. P. Vasilakos, Front. Immunol. 2013, 4, 230.

[28] R. B. Schnabel, J. Baumert, M. Barbalic, J. Dupuis, P. T. Ellinor, P. Durda, A. Dehghan, J. C. Bis, T. Illig, A. C. Morrison, N. S. Jenny, J. F. Keaney, Jr., C. Gieger, C. Tilley, J. F. Yamamoto, N. Khuseyinova, G. Heiss, M. Doyle, S. Blankenberg, C. Herder, J. D. Walston, Y. Zhu, R. S. Vasan, N. Klopp, E. Boerwinkle, M. G. Larson, B. M. Psaty, A. Peters, C. M. Ballantyne, J. C. Witteman, R. C. Hoogeveen, E. J. Benjamin, W. Koenig, R. P. Tracy, Blood 2010, 115, 5289.

[29] S. Thamphiwatana, P. Angsantikul, T. Escajadillo, Q. Zhang, J. Olson, B. T. Luk, S. Zhang, R. H. Fang, W. Gao, V. Nizet, L. Zhang, Proc. Natl. Acad. Sci. U.S.A. 2017, 114, 11488.

[30] B. T. Luk, C.-M. Jack Hu, R. H. Fang, D. Dehaini, C. Carpenter, W. Gao, L. Zhang, Nanoscale 2014, 6, 2730.

[31] J. Canton, D. Neculai, S. Grinstein, Nat. Rev. Immunol. 2013, 13, 621.

[32] K. Elvevold, J. Simon-Santamaria, H. Hasvold, P. McCourt, B. Smedsrød, K.K. Sørensen, Hepatology 2008, 48, 2007.

[33] I. Malovic, K. K. Sørensen, K. H. Elvevold, G. I. Nedredal, S. Paulsen, A. V. Erofeev, B. H. Smedsrød, P. A. McCourt, Hepatology 2007, 45, 1454.

[34] P. Pawar, P. K. Shin, S. A. Mousa, J. M. Ross, K. Konstantopoulos, J. Immunol. 2004, 173, 1258.

[35] J. Baum, R. H. Ward, D. J. Conway, Mol. Biol. Evol. 2002, 19, 223.

[36] D. E. Daigneault, K. L. Hartshorn, L. S. Liou, G. M. Abbruzzi, M. R. White, S.-K. Oh, A. I. Tauber, Blood 1992, 80, 3227; A. M. Gabali, J. J. Anzinger, G. T. Spear, L. L. Thomas, J. Virol. 2004, 78, 10833; K. Krebs, N. Böttinger, L. R. Huang, M. Chmielewski, S. Arzberger, G. Gasteiger, C. Jäger, E. Schmitt, F. Bohne, M. Aichler, W. Uckert, H. Abken, M. Heikenwalder, P. Knolle, U. Protzer, Gastroenterology 2013, 145, 456; O. Martinez, J. C. Johnson, A. Honko, B. Yen, R. S. Shabman, L. E. Hensley, G. G. Olinger, C. F. Basler, J Virol 2013, 87, 3801; D. Michlmayr, P. Andrade, K. Gonzalez, A. Balmaseda, E. Harris, Nat. Microbiol. 2017, 2, 1462.

[37] G. Zhang, G. R. Campbell, Q. Zhang, E. Maule, J. Hanna, W. Gao, L. Zhang, S. A. Spector, N. Chomont, N. Pujol, mBio 2020, 11, e00903.

[38] D. J. Anstee, Vox Sang. 1990, 58, 1; R. Khera, N. Das, Mol. Immunol. 2009, 46, 761.

[39] T. Kawasaki, T. Kawai, Front. Immunol. 2014, 5.

[40] O. Mandelboim, N. Lieberman, M. Lev, L. Paul, T. I. Arnon, Y. Bushkin, D. M. Davis, J. L. Strominger, J. W. Yewdell, A. Porgador, Nature 2001, 409, 1055.

[41] J. Lu, P. N. Tay, O. L. Kon, K. B. M. Reid, Biochem. J. 1996, 313, 473; C. Teh, Y. Le, S. H. Lee, J. Lu, Immunology 2000, 101, 225; Y. Liu, Y. Endo, D. Iwaki, M. Nakata, M. Matsushita, I. Wada, K. Inoue, M. Munakata, T. Fujita, J. Immunol. 2005, 175, 3150.

[42] A.-L. Favier, E. Gout, O. Reynard, O. Ferraris, J.-P. Kleman, V. Volchkov, C. Peyrefitte, N. M. Thielens, S. López, J. Virol. 2016, 90, 5256; J. S. McLellan, W. C. Ray, M. E. Peeples, Curr. Top. Microbiol. Immunol. 2013, 372, 83.

[43] H. Stoiber, I. Frank, M. Spruth, M. Schwendinger, B. Mullauer, J. M. Windisch, R. Schneider, H. Katinger, I. Ando, M. P. Dierich, Mol. Immunol. 1997, 34, 855.

[44] A. Lecube, G. Pachón, J. Petriz, C. Hernández, R. Simó, PLoS One 2011, 6, e23366.

[45] C. J. Van Oss, Infect. Immun. 1971, 4, 54.

[46] S. Hansen, S. Thiel, A. Willis, U. Holmskov, J. C. Jensenius, J Immunol 2000, 164, 2610.

[47] M. Stravalaci, I. Pagani, E. M. Paraboschi, M. Pedotti, A. Doni, F. Scavello, S. N. Mapelli, M. Sironi, L. Varani, M. Matkovic, A. Cavalli, D. Cesana, P. Gallina, N. Pedemonte, V. Capurro, N. Clementi, N. Mancini, P. Invernizzi, R. Rappuoli, S. Duga, B. Bottazzi, M. Uguccioni, R. Asselta, E. Vicenzi, A. Mantovani, C. Garlanda, medRxiv 2021, 2021.06.07.21258350.

[48] Y. K. Hahn, D. Hong, J. H. Kang, S. Choi, Micromachines 2016, 7, 139.

[49] S. H. Jung, Y. K. Hahn, S. Oh, S. Kwon, E. Um, S. Choi, J. H. Kang, Small 2018, 14, 1801731.

[50] J. H. Kang, H. Driscoll, M. Super, D. E. Ingber, Appl. Phys. Lett. 2016, 108, 213702; B. H. Jang, S. Kwon, J. H. Kang, Lab Chip 2019, 19, 2356.

[51] P. D. Stapleton, P. W. Taylor, Sci. Prog. 2002, 85, 57; A. Okano, N. A. Isley, D. L. Boger, Chem. Rev. 2017, 117, 11952.

[52] V. L. Thomas, B. A. Sanford, M. A. Ramsay, Microbiology 1993, 139, 623; J. Shuter, V. B. Hatcher, F. D. Lowy, Infect. Immun. 1996, 64, 310; C. J. Hackbarth, T. Kocagoz, S. Kocagoz, H. F. Chambers, Antimicrob. Agents Chemother. 1995, 39, 103.

[53] N. V. Prasadarao, C. A. Wass, K. S. Kim, Infect. Immun. 1997, 65, 2852; X. Tian, X. Zheng, Y. Sun, R. Fang, S. Zhang, X. Zhang, J. Lin, J. Cao, T. Zhou, Infect Drug Resist 2020, 13, 501; S. M. Khalifa, A. M. Abd El-Aziz, R. Hassan, E. S. Abdelmegeed, PLoS One 2021, 16, e0251594.

[54] M. Schindler, M. J. Osborn, D. E. Koppel, Nature 1980, 285, 261; Y.-C. Kim, Korean J. Chem. Eng. 1996, 13, 282.

[55] S. M. Opal, Contrib. Nephrol. 2010, 167, 14; T. Tanaka, M. Narazaki, T. Kishimoto, Cold Spring Harb. Perspect. Biol. 2014, 6, a016295.

[56] J.-S. Park, S.-J. Kim, S.-W. Lee, E.-J. Lee, K.-S. Han, S.-W. Moon, Y.-S. Hong, J. Emerg. Med. 2015, 49, 261.

[57] N. Brouwer, K. M. Dolman, M. van Houdt, M. Sta, D. Roos, T. W. Kuijpers, J. Immunol. 2008, 180, 4124.

[58] P. Lepper, T. Held, E. Schneider, E. Bölke, H. Gerlach, M. Trautmann, Intensive Care Med. 2002, 28, 824.

[59] R. S. Hotchkiss, K. C. Chang, M. H. Grayson, K. W. Tinsley, B. S. Dunne, C. G. Davis, D. F. Osborne, I. E. Karl, Proc. Natl. Acad. Sci. U.S.A. 2003, 100, 6724; R. S. Hotchkiss, S. B. Osmon, K. C. Chang, T. H. Wagner, C. M. Coopersmith, I. E. Karl, J. Immunol. 2005, 174, 5110.

[60] T. A. M. Claushuis, L. A. van Vught, B. P. Scicluna, M. A. Wiewel, P. M. C. Klein Klouwenberg, A. J. Hoogendijk, D. S. Y. Ong, O. L. Cremer, J. Horn, M. Franitza, M. R. Toliat, P. Nürnberg, A. H. Zwinderman, M. J. Bonten, M. J. Schultz, T. van der Poll, o. b. o. t. M. Diagnosis, R. S. o. S. Consortium, Blood 2016, 127, 3062; M. Gawaz, S. Vogel, Blood 2013, 122, 2550.

[61] S. Wang, R. Song, Z. Wang, Z. Jing, S. Wang, J. Ma, Front. Immunol. 2018, 9.

[62] B. P. Scicluna, F. Uhel, L. A. van Vught, M. A. Wiewel, A. J. Hoogendijk, I. Baessman, M. Franitza, P. Nürnberg, J. Horn, O. L. Cremer, M. J. Bonten, M. J. Schultz, T. van der Poll, Elife 2020, 9.

[63] M. Maier, E. V. Geiger, D. Henrich, C. Bendt, S. Wutzler, M. Lehnert, I. Marzi, Mol. Med. 2009, 15, 384.

[64] M. Garzón-Tituaña, M. A. Arias, J.L. Sierra-Monzón, E. Morte-Romea, L. Santiago, A. Ramirez-Labrada, L. Martinez-Lostao, J.R. Paño-Pardo, E. M. Galvez, J. Pardo, Front. Immunol. 2020, 11.

[65] K. N. Kangelaris, R. Clemens, X. Fang, A. Jauregui, T. Liu, K. Vessel, T. Deiss, P. Sinha, A. Leligdowicz, K. D. Liu, H. Zhuo, M. N. Alder, H. R. Wong, C. S. Calfee, C. Lowell, M. A. Matthay, Am. J. Physiol. Lung Cell. Mol. Physiol. 2021, 320, L892.

[66] D. Braga, M. Barcella, F. D’Avila, S. Lupoli, F. Tagliaferri, M. H. Santamaria, F. A. DeLano, G. Baselli, G.W. Schmid-Schönbein, E. B. Kistler, F. Aletti, C. Barlassina, Exp. Biol. Med. (Maywood) 2017, 242, 1462.

[67] I. N. Shalova, J. Y. Lim, M. Chittezhath, A. S. Zinkernagel, F. Beasley, E. Hernández-Jiménez, V. Toledano, C. Cubillos-Zapata, A. Rapisarda, J. Chen, K. Duan, H. Yang, M. Poidinger, G. Melillo, V. Nizet, F. Arnalich, E. López-Collazo, S. K. Biswas, Immunity 2015, 42, 484.

[68] J. Ma, C. Chen, A. S. Barth, C. Cheadle, X. Guan, L. Gao, Mediat. Inflamm. 2015, 2015, 984825; L. Bu, Z. W. Wang, S. Q. Hu, W. J. Zhao, X. J. Geng, T. Zhou, L. Zhuo, X. B. Chen, Y. Sun, Y. L. Wang, X. M. Li, Comput. Math. Methods Med. 2020, 2020, 8741739.

[69] V. M. Ranieri, B. T. Thompson, P. S. Barie, J. F. Dhainaut, I. S. Douglas, S. Finfer, B. Gårdlund, J. C. Marshall, A. Rhodes, A. Artigas, D. Payen, J. Tenhunen, H. R. Al-Khalidi, V. Thompson, J. Janes, W. L. Macias, B. Vangerow, M. D. Williams, N. Engl. J. Med. 2012, 366, 2055.

[70] I. K. Herrmann, M. Urner, F. M. Koehler, M. Hasler, B. Roth-Z’Graggen, R. N. Grass, U. Ziegler, B. Beck-Schimmer, W. J. Stark, Small 2010, 6, 1388.

[71] L. Rao, L.-L. Bu, J.-H. Xu, B. Cai, G.-T. Yu, X. Yu, Z. He, Q. Huang, A. Li, S.-S. Guo, W.-F. Zhang, W. Liu, Z.-J. Sun, H. Wang, T.-H. Wang, X.-Z. Zhao, Small 2015, 11, 6225.

[72] C. Garcia-Vidal, G. Sanjuan, E. Moreno-García, P. Puerta-Alcalde, N. Garcia-Pouton, M. Chumbita, M. Fernandez-Pittol, C. Pitart, A. Inciarte, M. Bodro, L. Morata, J. Ambrosioni, Grafia, F. Meira, I. Macaya, C. Cardozo, C. Casals, A. Tellez, P. Castro, F. Marco, F. García Mensa, J. A. Martínez, A. Soriano, V. Rico, M. Hernández-Meneses, D. Agüero, B. Torres, A. González, L. de la Mora, J. Rojas, L. Linares, B. Fidalgo, N. Rodriguez, D. Nicolas, L. Albiach, J. Muñoz, A. Almuedo, D. Camprubí, M. Angeles Marcos, D. Camprubí, C. Cilloniz, S. Fernández, J. M. Nicolas, A. Torres, Clin. Microbiol. Infect. 2021, 27, 83.

[73] P. Avirutnan, R. E. Hauhart, M. A. Marovich, P. Garred, J. P. Atkinson, M. S. Diamond, mBio 2011, 2, e00276.

[74] N. Inanc, G. Mumcu, E. Birtas, Y. Elbir, S. Yavuz, T. Ergun, I. Fresko, H. Direskeneli, Rheumatol. 2005, 32, 287.

[75] Y. Wu, X. Huang, J. Sun, T. Xie, Y. Lei, J. Muhammad, X. Li, X. Zeng, F. Zhou, H. Qin, L. Shao, Q. Zhang, mSphere 2020, 5, e00362.

[76] W. He, S. Neil, H. Kulkarni, E. Wright, B. K. Agan, V. C. Marconi, M. J. Dolan, R. A. Weiss, S. K. Ahuja, Cell Host Microbe 2008, 4, 52.

[77] AIDS Res. Hum. Retrovir. 2004, 20, 327.

[78] E. Jaskiewicz, M. Jodłowska, R. Kaczmarek, A. Zerka, Parasit. Vectors 2019, 12, 317.

[79] W.-H. Tham, D. W. Wilson, S. Lopaticki, C. Q. Schmidt, P. B. Tetteh-Quarcoo, P. N. Barlow, D. Richard, J. E. Corbin, J. G. Beeson, A. F. Cowman, Proc. Natl. Acad. Sci. U.S.A. 2010, 201008151.

[80] S. Hojyo, M. Uchida, K. Tanaka, R. Hasebe, Y. Tanaka, M. Murakami, T. Hirano, Inflamm. Regen. 2020, 40, 37; R. J. Jose, A. Manuel, Lancet Respir. Med. 2020, 8, e46.

[81] R. S. Hotchkiss, S. B. Osmon, K. C. Chang, T. H. Wagner, C. M. Coopersmith, I. E. Karl, J. Immunol. 2005, 174, 5110.

[82] T. A. M. Claushuis, L. A. van Vught, B. P. Scicluna, M. A. Wiewel, P. M. C. Klein Klouwenberg, A. J. Hoogendijk, D. S. Y. Ong, O. L. Cremer, J. Horn, M. Franitza, M. R. Toliat, P. Nürnberg, A. H. Zwinderman, M. J. Bonten, M. J. Schultz, T. van der Poll, o. b. o. t. M. Diagnosis, R. S. o. S. Consortium, Blood 2016, 127, 3062.

[83] M. Gawaz, S. Vogel, Blood 2013, 122, 2550.

[84] S. Wang, R. Song, Z. Wang, Z. Jing, S. Wang, J. Ma, Front. Immunol. 2018, 9.

[85] B. P. Scicluna, F. Uhel, L. A. van Vught, M. A. Wiewel, A. J. Hoogendijk, I. Baessman, M. Franitza, P. Nürnberg, J. Horn, O. L. Cremer, M. J. Bonten, M. J. Schultz, T. van der Poll, Elife 2020, 9.

[86] M. Maier, E. V. Geiger, D. Henrich, C. Bendt, S. Wutzler, M. Lehnert, I. Marzi, Mol. Med. 2009, 15, 384.

[87] M. Garzón-Tituaña, M. A. Arias, J.L. Sierra-Monzón, E. Morte-Romea, L. Santiago, A. Ramirez-Labrada, L. Martinez-Lostao, J.R. Paño-Pardo, E. M. Galvez, J. Pardo, Front. Immunol. 2020, 11.

[88] K. N. Kangelaris, R. Clemens, X. Fang, A. Jauregui, T. Liu, K. Vessel, T. Deiss, P. Sinha, A. Leligdowicz, K. D. Liu, H. Zhuo, M. N. Alder, H. R. Wong, C. S. Calfee, C. Lowell, M. A. Matthay, Am. J. Physiol. Lung Cell. Mol. Physiol. 2021, 320, L892.

[89] D. Braga, M. Barcella, F. D’Avila, S. Lupoli, F. Tagliaferri, M. H. Santamaria, F. A. DeLano, G. Baselli, G. W. Schmid-Schönbein, E. B. Kistler, F. Aletti, C. Barlassina, Exp. Biol. Med. (Maywood) 2017, 242, 1462.

[90] I. N. Shalova, J. Y. Lim, M. Chittezhath, A. S. Zinkernagel, F. Beasley, E. Hernández-Jiménez, V. Toledano, C. Cubillos-Zapata, A. Rapisarda, J. Chen, K. Duan, H. Yang, M. Poidinger, G. Melillo, V. Nizet, F. Arnalich, E. López-Collazo, S. K. Biswas, Immunity 2015, 42, 484.

[91] J. Ma, C. Chen, A. S. Barth, C. Cheadle, X. Guan, L. Gao, Mediat. Inflamm. 2015, 2015, 984825.

[92] L. Bu, Z. W. Wang, S. Q. Hu, W. J. Zhao, X. J. Geng, T. Zhou, L. Zhuo, X. B. Chen, Y. Sun, Y. L. Wang, X. M. Li, Comput. Math. Methods Med. 2020, 2020, 8741739.

[93] V. M. Ranieri, B. T. Thompson, P. S. Barie, J. F. Dhainaut, I. S. Douglas, S. Finfer, B. Gårdlund, J. C. Marshall, A. Rhodes, A. Artigas, D. Payen, J. Tenhunen, H. R. Al-Khalidi, V. Thompson, J. Janes, W. L. Macias, B. Vangerow, M. D. Williams, N. Engl. J. Med. 2012, 366, 2055.

[94] I. K. Herrmann, M. Urner, F. M. Koehler, M. Hasler, B. Roth-Z’Graggen, R. N. Grass, U. Ziegler, B. Beck-Schimmer, W. J. Stark, Small 2010, 6, 1388.

[95] L. Rao, L.-L. Bu, J.-H. Xu, B. Cai, G.-T. Yu, X. Yu, Z. He, Q. Huang, A. Li, S.-S. Guo, W.-F. Zhang, W. Liu, Z.-J. Sun, H. Wang, T.-H. Wang, X.-Z. Zhao, Small 2015, 11, 6225.

[96] C. Garcia-Vidal, G. Sanjuan, E. Moreno-García, P. Puerta-Alcalde, N. Garcia-Pouton, M. Chumbita, M. Fernandez-Pittol, C. Pitart, A. Inciarte, M. Bodro, L. Morata, J. Ambrosioni, Grafia, F. Meira, I. Macaya, C. Cardozo, C. Casals, A. Tellez, P. Castro, F. Marco, F. García, J. Mensa, J. A. Martínez, A. Soriano, V. Rico, M. Hernández-Meneses, D. Agüero, B. Torres, A. González, L. de la Mora, J. Rojas, L. Linares, B. Fidalgo, N. Rodriguez, D. Nicolas, L. Albiach, J. Muñoz, A. Almuedo, D. Camprubí, M. Angeles Marcos, D. Camprubí, C. Cilloniz, S. Fernández, J. M. Nicolas, A. Torres, Clin. Microbiol. Infect. 2021, 27, 83.

[97] P. Avirutnan, R. E. Hauhart, M. A. Marovich, P. Garred, J. P. Atkinson, M. S. Diamond, mBio 2011, 2, e00276.

[98] N. Inanc, G. Mumcu, E. Birtas, Y. Elbir, S. Yavuz, T. Ergun, I. Fresko, H. Direskeneli, J. Rheumatol. 2005, 32, 287.

[99] Y. Wu, X. Huang, J. Sun, T. Xie, Y. Lei, J. Muhammad, X. Li, X. Zeng, F. Zhou, H. Qin, L. Shao, Q. Zhang, mSphere 2020, 5, e00362.

[100] W. He, S. Neil, H. Kulkarni, E. Wright, B. K. Agan, V. C. Marconi, M. J. Dolan, R. A. Weiss, S. K. Ahuja, Cell Host Microbe 2008, 4, 52.

[101] H. Ying, X. Ji, M. L. Hart, K. Gupta, M. Saifuddin, M. R. Zariffard, G. T. Spear, AIDS Res. Hum. Retrovir. 2004, 20, 327.

[102] E. Jaskiewicz, M. Jodłowska, R. Kaczmarek, A. Zerka, Parasit. Vectors 2019, 12, 317.

[103] W.-H. Tham, D. W. Wilson, S. Lopaticki, C. Q. Schmidt, P. B. Tetteh-Quarcoo, P. N. Barlow, D. Richard, J. E. Corbin, J. G. Beeson, A. F. Cowman, Proc. Natl. Acad. Sci. U.S.A. 2010, 201008151.

[104] S. Hojyo, M. Uchida, K. Tanaka, R. Hasebe, Y. Tanaka, M. Murakami, T. Hirano, Inflamm. Regen. 2020, 40, 37.

[105] R. J. Jose, A. Manuel, Lancet Respir. Med. 2020, 8, e46

