## Supporting Information for "Human cell-camouflaged nanomagnetic scavengers restore immune homeostasis in a rodent model with bacteremia"

(UNIST), UNIST gil 50, Ulsan, Republic of Korea;

^*^Corresponding Authors:

Joo H. Kang, Ph.D.

Department of Biomedical Engineering,

Ulsan National Institute of Science and Technology (UNIST),

Ulsan National Institute of Science and Technology (UNIST), UNIST gil 50, Ulju gun, Ulsan, Republic of Korea

^†^ These authors contributed equally.

^†^ These authors contributed equally.

This PDF file includes

Figs. S1 to S6

Table S1

Legends for tables S2 to S4

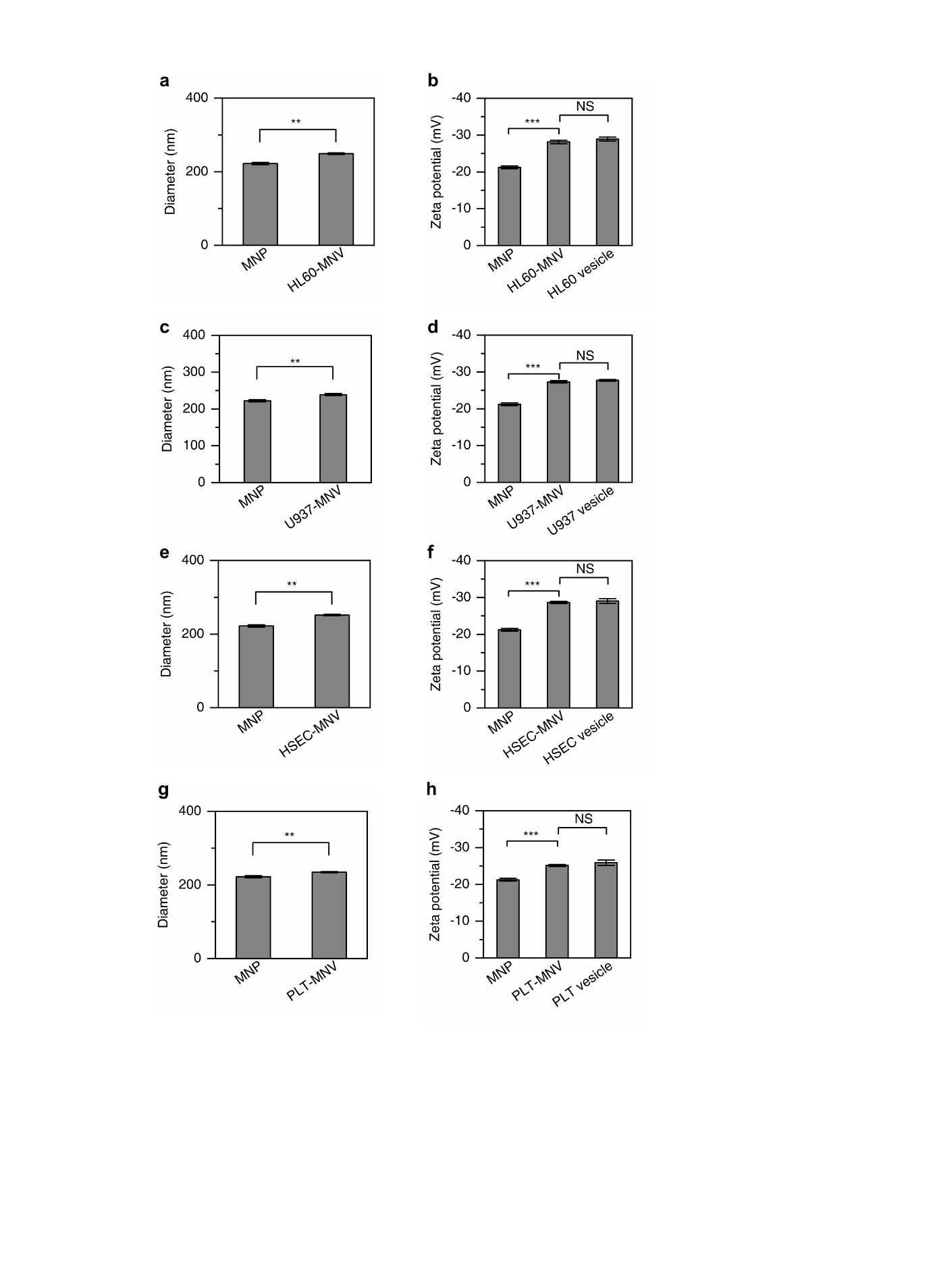

**Figure S1.** Characterization of the MNVs made of several cell membranes. a, b) Hydrodynamic diameter a) and Zeta potential b) of MNPs, MNVs, and membrane-vesicles without cores made of HL60. c,d) Hydrodynamic diameter c) and Zeta potential d) of MNPs, MNVs, and membrane-vesicles without cores made of U937. e, f) Hydrodynamic diameter e) and Zeta potential f) of MNPs, MNVs, and membrane-vesicles without cores made of hHSEC. g, h) Hydrodynamic diameter g) and Zeta potential h) of MNPs, MNVs, and membrane-vesicles without cores made of hPLT. (*n*=3). Data were expressed as means ± SEM. Statistical significance was calculated by a two-tailed Student’s t test. ***P* < 0.005; ****P* < 0.001; NS, not significant.

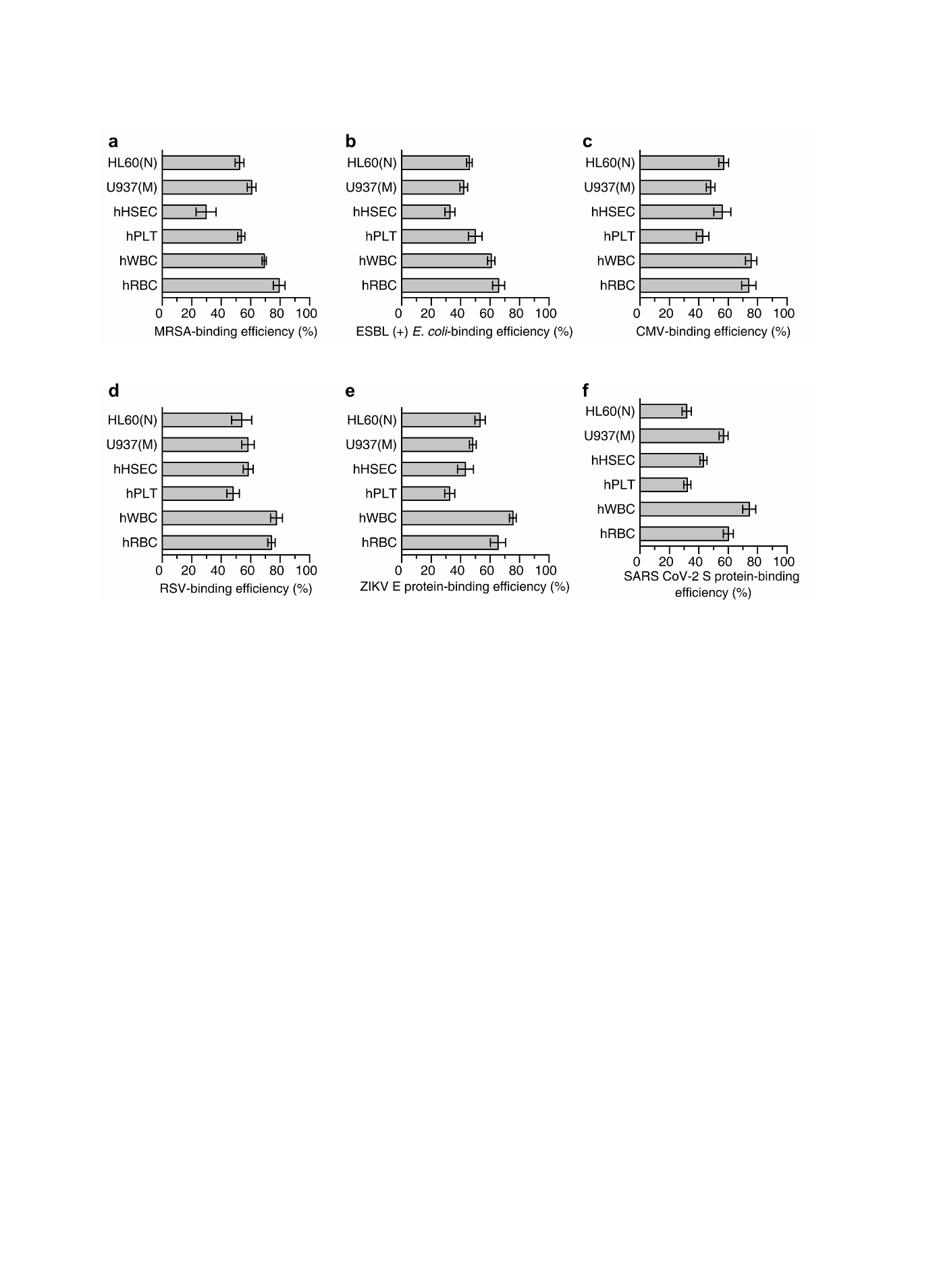

**Figure S2.** Binding efficiency of the MNVs made of several cell membranes to the pathogens. The binding efficiencies of the MNVs made of HL60, U937, hHSEC, hPLT, hWBC, and hRBC membranes to a) MRSA , b) ESBL-EC , c) CMV, d) RSV, e) Zika virus E protein, and f) SARS-CoV-2 spike protein. (*n*=3). Data were expressed as means ± SEM.

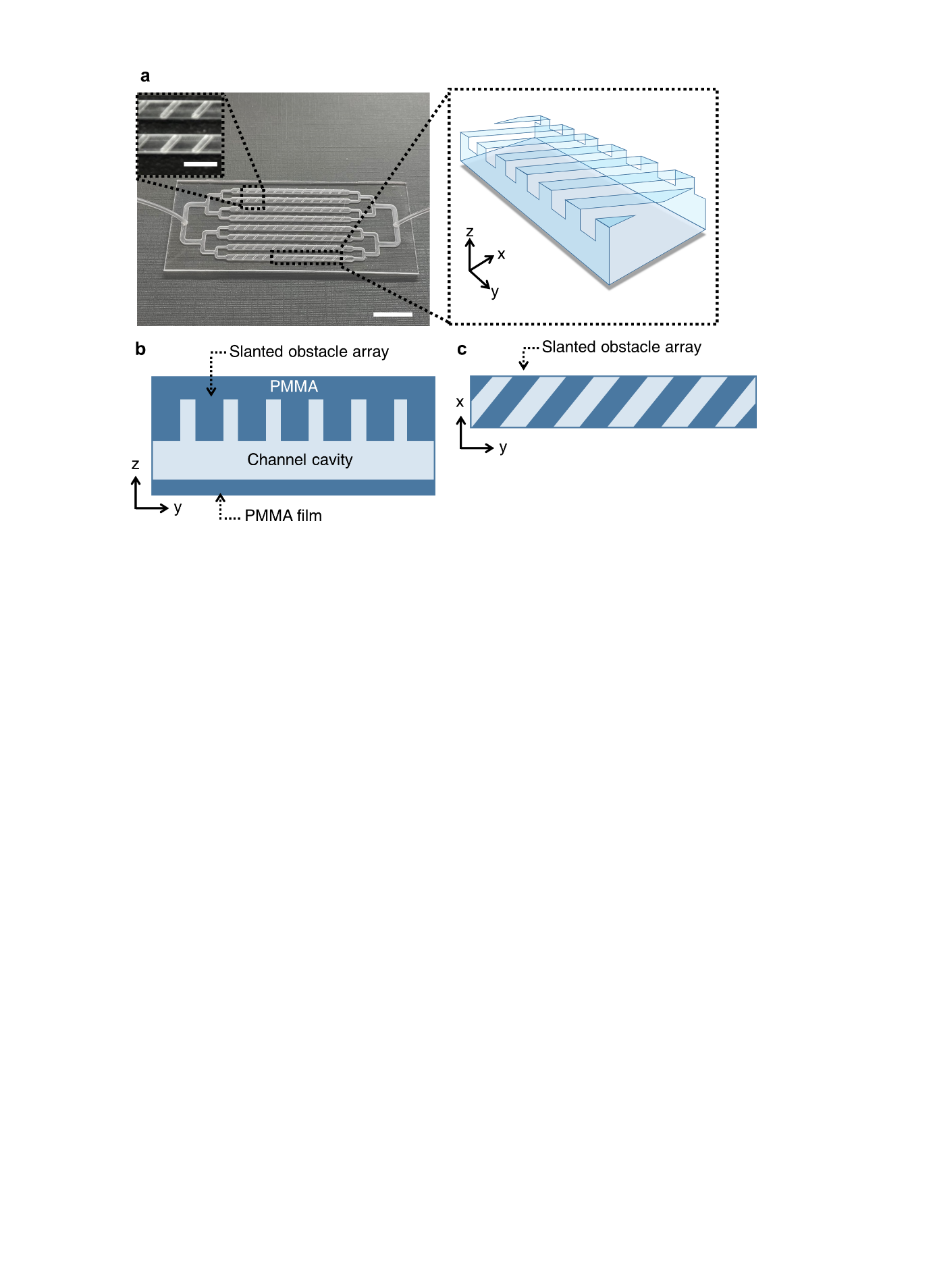

**Figure S3.** Photograph of the microfluidic chip in the blood-cleansing device. a) A representative photograph of the microfluidic chip used in the blood-cleansing device. Scale bar, 2 cm. The inset image shows slanted obstacles patterned on top of the channel in the device. Scale bar, 0.5 cm. b) The side view and c) top view of the singl channel of the blood cleansing device

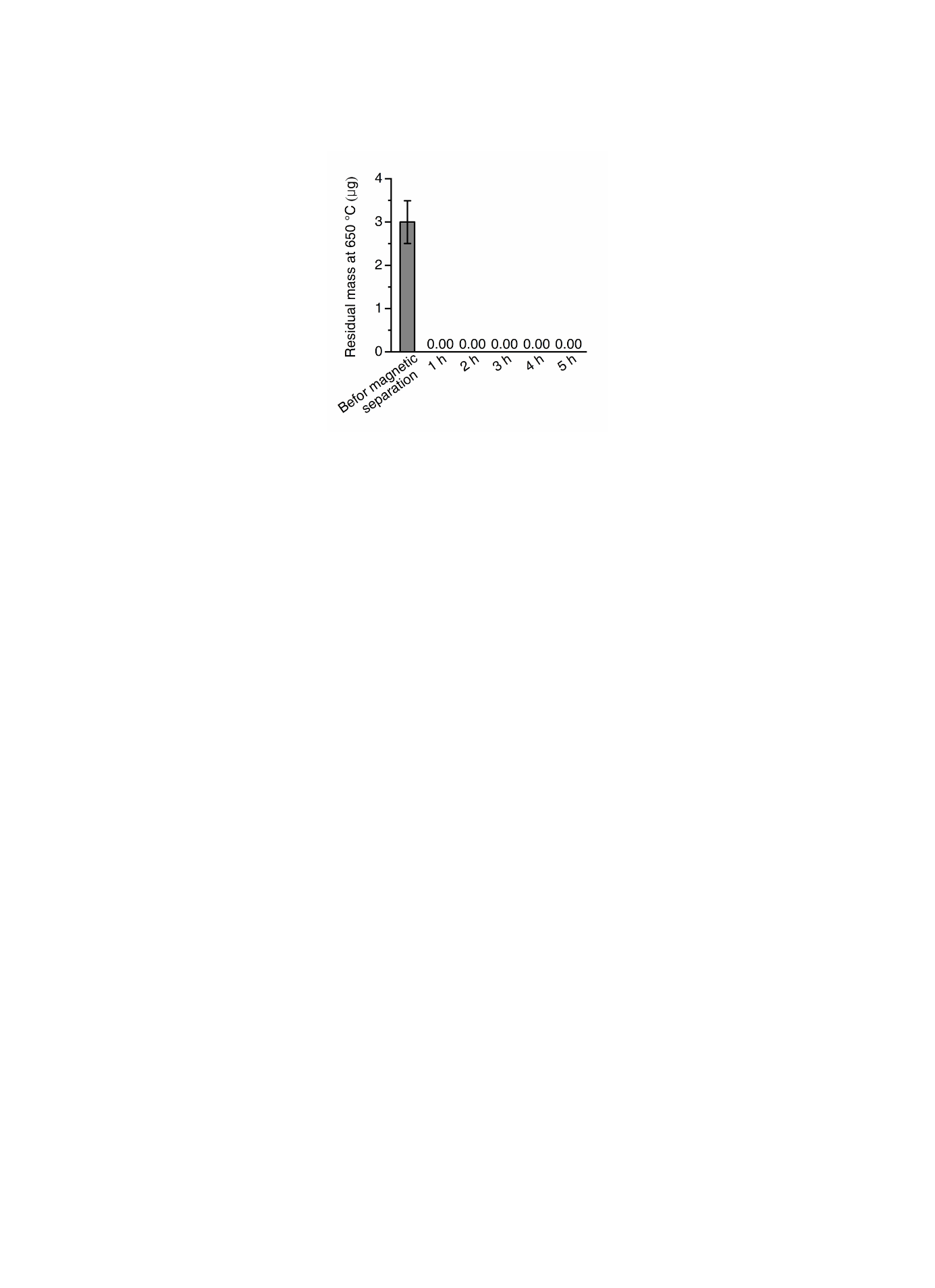

**Figure S4.** Removal of MNVs from the flowing blood using the magnetic separation device. The residual mass of the hRBC-MNVs in the blood samples containing 24 μg mL^-1^ hRBC-MNVs measured by TGA after passing through the extracorporeal magnetic separation device. 10 mL of the MNVs-containing blood sample was circulated through the magnetic separation device at a flow rate of 10 mL h^-1^ and 100 μL of blood was collected every hour (~5 h) at the outlet of the device. The results indicate that MNVs were completely removed by the magnetic separation device. Data were expressed as means ± SEM.

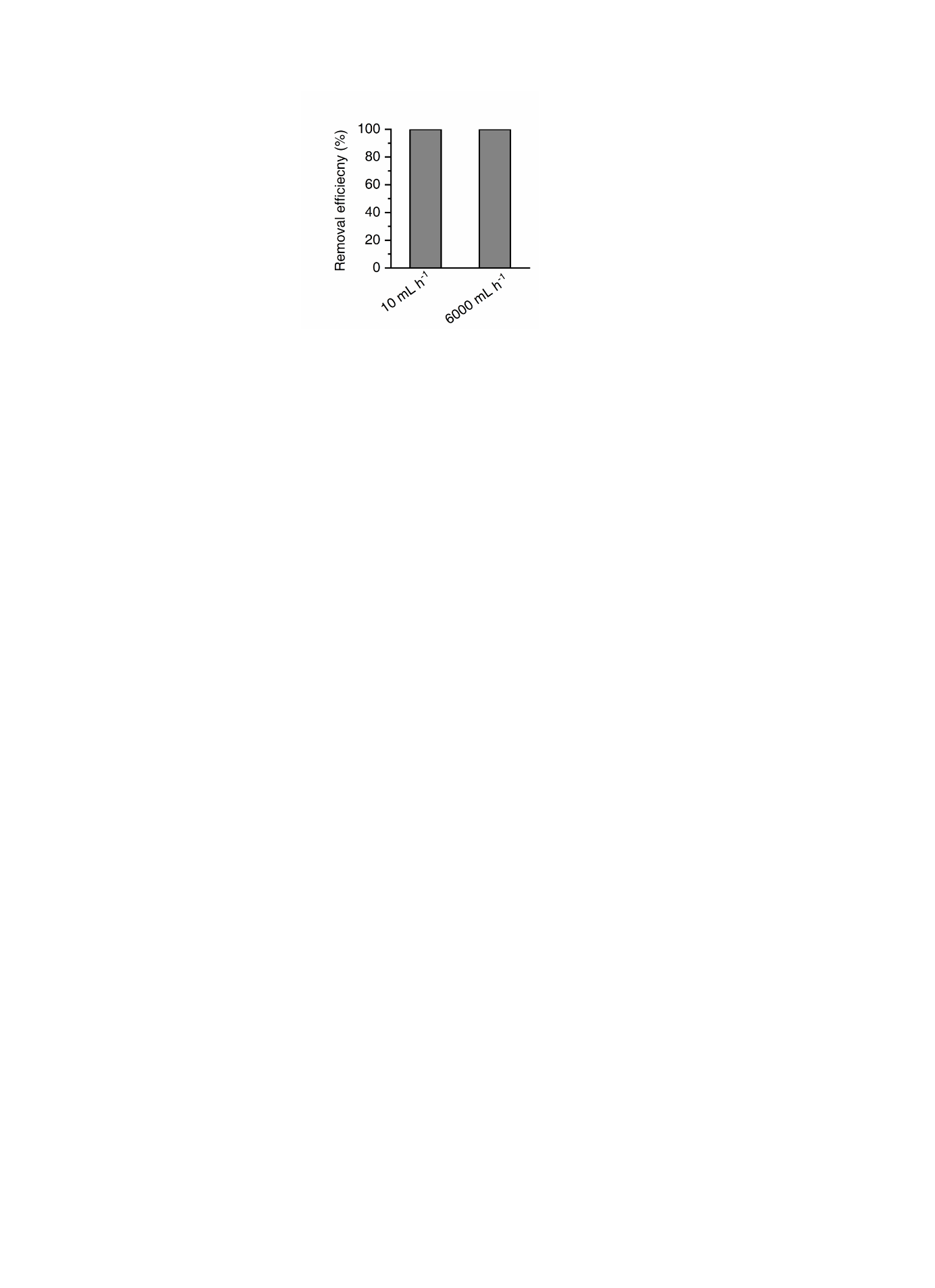

**Figure S5.** The magnetic removal efficiency of MRSA-bound hRBC-MNVs using the blood cleansing device with hRBC-MNVs at different flow rates. The removal efficiency of MRSA-bound hRBC-MNVs from the blood was measured after a single passing through the magnetic separation device at flow rates of 10 mL h^-1^ and 6000 mL h^-1^. The later flow rate is clinically used in extracorporeal hemoperfusion. The initial CFU of the MRSA bounded to hRBC-MNVs was measured as 10^4^ CFU mL^-1^ in the blood before passing through the device. Data were expressed as means ± SEM.

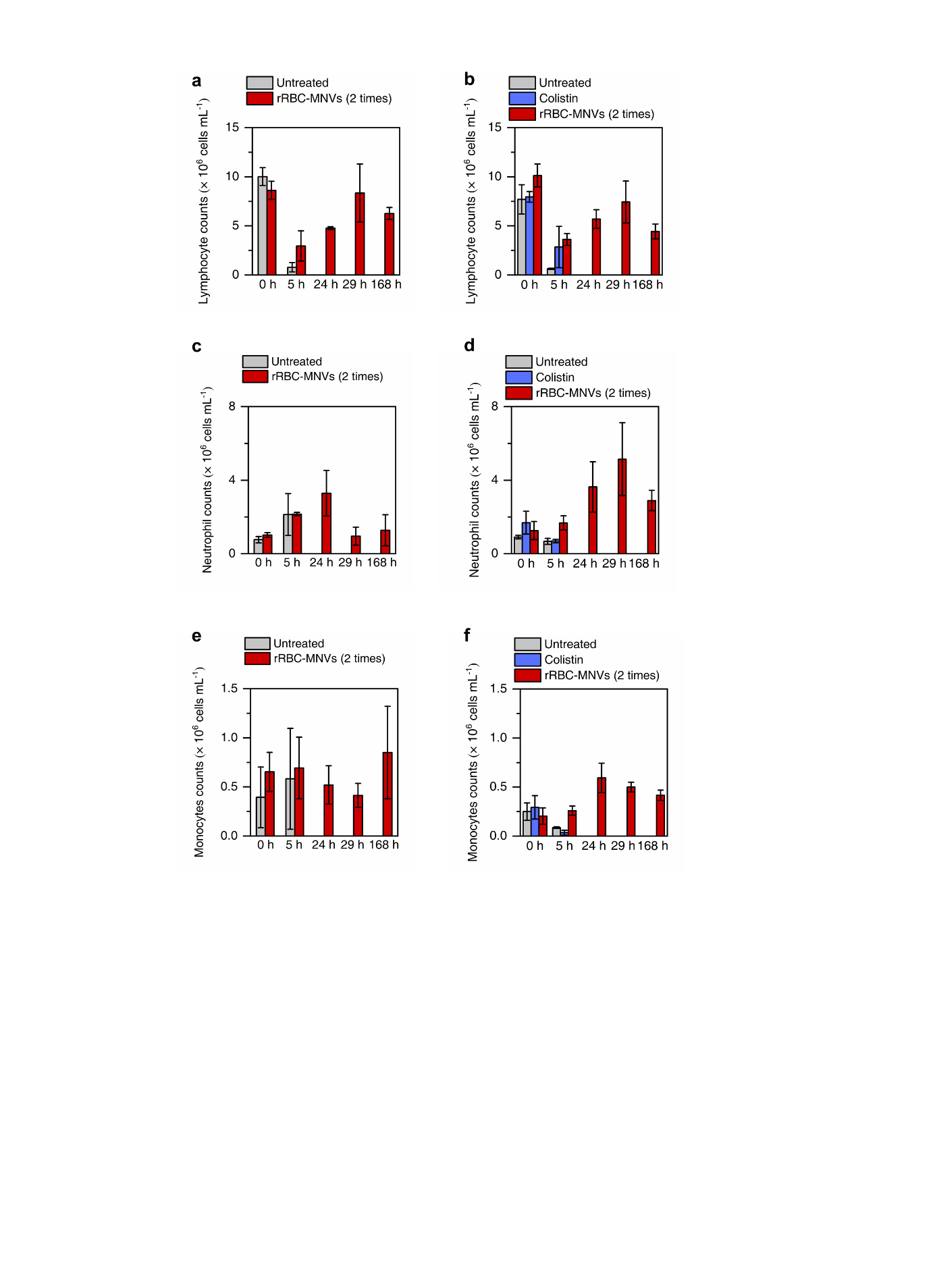

**Figure S6.** Major WBCs counts in bacteremic rats with or without blood-cleansing treatment using rRBC-MNVs. a) Lymphocyte counts in MRSA-infected rats with two rounds of 5-h treatment using rRBC-MNVs and without treatment. b) Lymphocyte counts in CR *E. coli*-infected rats with two rounds of 5-h treatment using rRBC-MNVs, colistin administration, and without treatment. c) Neutrophil counts in MRSA-infected rats with two rounds of 5-h treatment using rRBC-MNVs and without treatment. d) Neutrophil counts in CR *E. coli*-infected rats with two rounds of 5-h treatment using rRBC-MNVs, colistin administration, and without treatment. e) Monocyte counts in MRSA-infected rats with two rounds of 5-h treatment using rRBC-MNVs and without treatment. f) Monocyte counts in CR *E. coli*-infected rats with two rounds of 5-h treatment using rRBC-MNVs, colistin administration, and without treatment. (*n*=3). Data were expressed as means ± SEM.

Removal efficiency: 100%

*Aerococcus vaginalis*

*Agathobaculum desmolans*

*Anaerobiospirillum succiniciproducens*

*Anaerobium acetethylicum*

*Anaerobutyricum hallii*

*Blautia luti*

*Blautia wexlerae*

*Brachyspira hampsonii*

*Buchananella hordeovulneris*

*Campylobacter upsaliensis*

*Candidatus Pelagibacter ubique*

*Clostridium paraputrificum*

*Coprococcus catus*

*Corynebacterium amycolatum*

*Corynebacterium auriscanis*

*Corynebacterium confusum*

*Corynebacterium lowii*

*Demequina aestuarii*

*Desulfovibrio simplex*

*Dietzia maris*

*Dorea formicigenerans*

*Dorea longicatena*

*Enterocloster clostridioformis*

*Enterococcus dispar*

*Enterococcus faecalis*

*Enterococcus gallinarum*

*Escherichia marmotae*

*Eubacterium ramulus*

*Frederiksenia canicola*

*Helicobacter bilis*

*Helicobacter canicola*

*Helicobacter canis*

*Helicobacter cinaedi*

*Helicobacter winghamensis*

*Hespellia porcina*

*Klebsiella pneumoniae*

*Klebsiella variicola*

*Lachnospira multipara*

*Longibaculum muris*

*Nitrososphaera viennensis*

*Paludibacter propionicigenes*

*Pasteurella stomatis*

*Phascolarctobacterium succinatutens*

*Pseudoflavonifractor phocaeensis*

*Staphylococcus felis*

*Staphylococcus intermedius*

*Staphylococcus simulans*

*Streptococcus canis*

*Synechococcus rubescens*

*Winkia neuii*

Removal efficiency: 99.9~99.99%

*Agathobaculum butyriciproducens*

*Alistipes putredinis*

*Alistipes shahii*

*Allobaculum stercoricanis*

*Amedibacillus dolichus*

*Anaerostipes hadrus*

*Anaerotignum faecicola*

*Bacteroides caccae*

*Bacteroides fragilis*

*Bacteroides koreensis*

*Bacteroides pyogenes*

*Bacteroides stercoris*

*Bacteroides thetaiotaomicron*

*Bacteroides uniformis*

*Bacteroides xylanisolvens*

*Bifidobacterium catenulatum*

*Blautia faecis*

*Blautia marasmi*

*Blautia obeum*

*Butyricicoccus pullicaecorum*

*Campylobacter showae*

*Caproiciproducens galactitolivorans*

*Clostridium methylpentosum*

*Clostridium spiroforme*

*Clostridium tertium*

*Dialister invisus*

*Erysipelatoclostridium ramosum*

*Escherichia fergusonii*

*Eubacterium rectale*

*Eubacterium ventriosum*

*Faecalibacterium prausnitzii*

*Faecalimonas umbilicata*

*Flavonifractor plautii*

*Flintibacter butyricus*

*Fournierella massiliensis*

*Fusicatenibacter saccharivorans*

*Fusobacterium mortiferum*

*Fusobacterium perfoetens*

*Gemmiger formicilis*

*Holdemania massiliensis*

*Hungatella xylanolytica*

*Kineothrix alysoides*

*Lachnoclostridium pacaense*

*Lachnospira eligens*

*Lachnospira pectinoschiza*

*Lactobacillus rogosae*

*Mediterranea massiliensis*

*Mediterraneibacter glycyrrhizinilyticus*

*Monoglobus pectinilyticus*

*Mucispirillum schaedleri*

*Oscillibacter ruminantium*

*Parabacteroides distasonis*

*Parabacteroides merdae*

*Paraprevotella clara*

*Peptococcus niger*

*Peptostreptococcus canis*

*Phocaeicola coprocola*

*Phocaeicola coprophilus*

*Phocaeicola plebeius*

*Phocaeicola vulgatus*

*Prevotella stercorea*

*Pseudomonas matsuisoli*

*Romboutsia sedimentorum*

*Roseburia faecis*

*Roseburia hominis*

*Roseburia intestinalis*

*Roseburia inulinivorans*

*Ruminococcus gnavus*

*Ruminococcus lactaris*

*Schaalia canis*

*Sutterella stercoricanis*

Other

Removal efficiency: 99.0~99.9%

*Blautia argi*

*Collinsella intestinalis*

*Eubacterium coprostanoligenes*

*Haemophilus haemoglobinophilus*

*Peptacetobacter hiranonis*

*Porphyromonas cangingivalis*

*Sutterella massiliensis*

*Turicibacter sanguinis*

*Tyzzerella nexilis*

Removal efficiency: 90.0~99.0%

*Blautia schinkii*

*Bacteroides cellulosilyticus*

*Catenibacterium mitsuokai*

*Holdemanella biformis*

*Megamonas funiformis*

**Table S1. The list of the pathogens in human fecal material inoculated in whole blood removed using hRBC-MNVs.**

**LEGENDS FOR SUPPLEMENTARY TABLES**

Table S2. List of the DEGs between blood-cleansing treated and untreated samples (separate file). DEGs between the CR *E. coli*-infected rats receiving two rounds of the 5-hour blood-cleansing treatment for two serial days (treated) and the CR *E. coli*-infected rats without treatment (untreated) were listed with gene ID, transcript ID, fold change, *P*-value (quasi-likelihood F test), and the read counts for the treated and untreated samples.

Table S3. List of the significantly up- and down-regulated DEGs (separate file). Significantly up- and down-regulated DEGs for treated and untreated samples are collected which have a log_2_ fold change value greater than 1.5 with a *P*-value of less than 0.05. The collected DEGs are listed with gene ID, transcript ID, fold change, P-value (quasi-likelihood F test), and the read counts for the treated and untreated samples.

Table S4. List of the significantly up- and down-regulated DEGs which are associated with sepsis (separate file). Sepsis-associated genes are selected from the significantly up- and down-regulated DEGs (P < 0.05 & log2 fold change > 1.5) between treated and untreated samples using the Open Targets Platform. The collected DEGs are listed with gene ID, transcript ID, fold change, *P*-value (quasi-likelihood F test), and the read counts for the treated and untreated samples.
